# Genetically informed precision drug repurposing for lung function and implications for respiratory infection

**DOI:** 10.1101/2020.06.25.20139816

**Authors:** William R. Reay, Sahar I. El Shair, Michael P. Geaghan, Carlos Riveros, Elizabeth G. Holliday, Mark A. McEvoy, Stephen Hancock, Roseanne Peel, Rodney J. Scott, John R. Attia, Murray J. Cairns

## Abstract

Impaired lung function is associated with significant morbidity and mortality. Restrictive and obstructive lung disorders are a large contributor to decreased lung function, as well as the acute impact of infection. Measures of pulmonary function are heritable, and thus, we sought to utilise genomics to propose novel drug repurposing candidates which could improve respiratory outcomes. Lung function measures were found to be genetically correlated with metabolic and hormone traits which could be pharmacologically modulated, with a causal effect of increased fasting glucose on diminished lung function supported by latent causal variable models and Mendelian randomisation. We developed polygenic scores for lung function specifically within pathways with known drug targets to prioritise individuals who may benefit from particular drug repurposing opportunities, accompanied by transcriptome-wide association studies to identify drug-gene interactions with potential lung function increasing modes of action. These drug repurposing candidates were further considered relative to the host-viral interactome of three viruses with associated respiratory pathology (SARS-CoV2, influenza, and human adenovirus). We uncovered an enrichment amongst glycaemic pathways of human proteins which putatively interact with virally expressed SARS-CoV2 proteins, suggesting that antihyperglycaemic agents may have a positive effect both on lung function and SARS-CoV2 progression.

## MAIN

Optimal lung (pulmonary) function is vital for the ongoing maintenance of homeostasis, with reduced pulmonary function associated with a marked increase in the risk of mortality^1,2^. This is particularly critical due to the considerable number of disorders for which diminished pulmonary function is a clinical hallmark. For instance, chronic obstructive pulmonary disease (COPD), characterised by an irreversible limitation of airflow, is one of the leading causes of death worldwide^3^. Pulmonary manifestations are also common amongst disorders not directly classified as respiratory conditions, including diabetes^4,5^, congenital heart disease^6^, and inflammatory bowel disease^7,8^. Bacterial and viral infection, such as *Streptococcus pneumoniae, Mycobacterium tuberculosis*, influenza, and coronaviruses, also cause severe declines in respiratory function. In order to better manage the spectrum of respiratory disorders there is a desperate need for new interventions, including those that can be targeted to an individual’s heterogeneous risk factors. While the development pathway for new compounds is difficult, there are likely to be opportunities for precision repurposing of existing drugs to enhance lung function and improve patient outcomes.

Spirometry measures of pulmonary function have been shown to display significant heritability both in twin designs and genome-wide association studies (GWAS)^9–11^. Genomics may reveal clinically relevant insights into the biology underlying lung function, and thus, could be leveraged for drug repurposing. We sought to interrogate the genomic architecture of three spirometry indices to propose drug repurposing candidates which could be used to improve lung function: forced expiratory volume in one second (FEV_1_), forced vital capacity (FVC), and their ratio (FEV_1_/FVC). Firstly, we assessed each lung function trait for evidence of genetic correlation with biochemical traits that could be pharmacologically modulated, followed by models to investigate whether there was evidence of causation. The previously developed *pharmagenic enrichment score* framework was then implemented to identify druggable pathways enriched with lung function associated variation and calculate pathway specific polygenic scores to prioritise individuals who may benefit from a repurposed compound which interacts with the pathway^12^. A transcriptome-wide association study of FEV_1_ and FVC was also undertaken to reveal genes which could be targeted by existing drugs that may increase pulmonary function. Finally, we considered the repurposing candidates proposed by these strategies in the context of three respiratory viruses (SARS-CoV2, influenza, and human adenovirus), specifically, analysing the interactions between viral and human proteins. An overview schematic of this study is detailed in figure 1.

**Figure 1.**
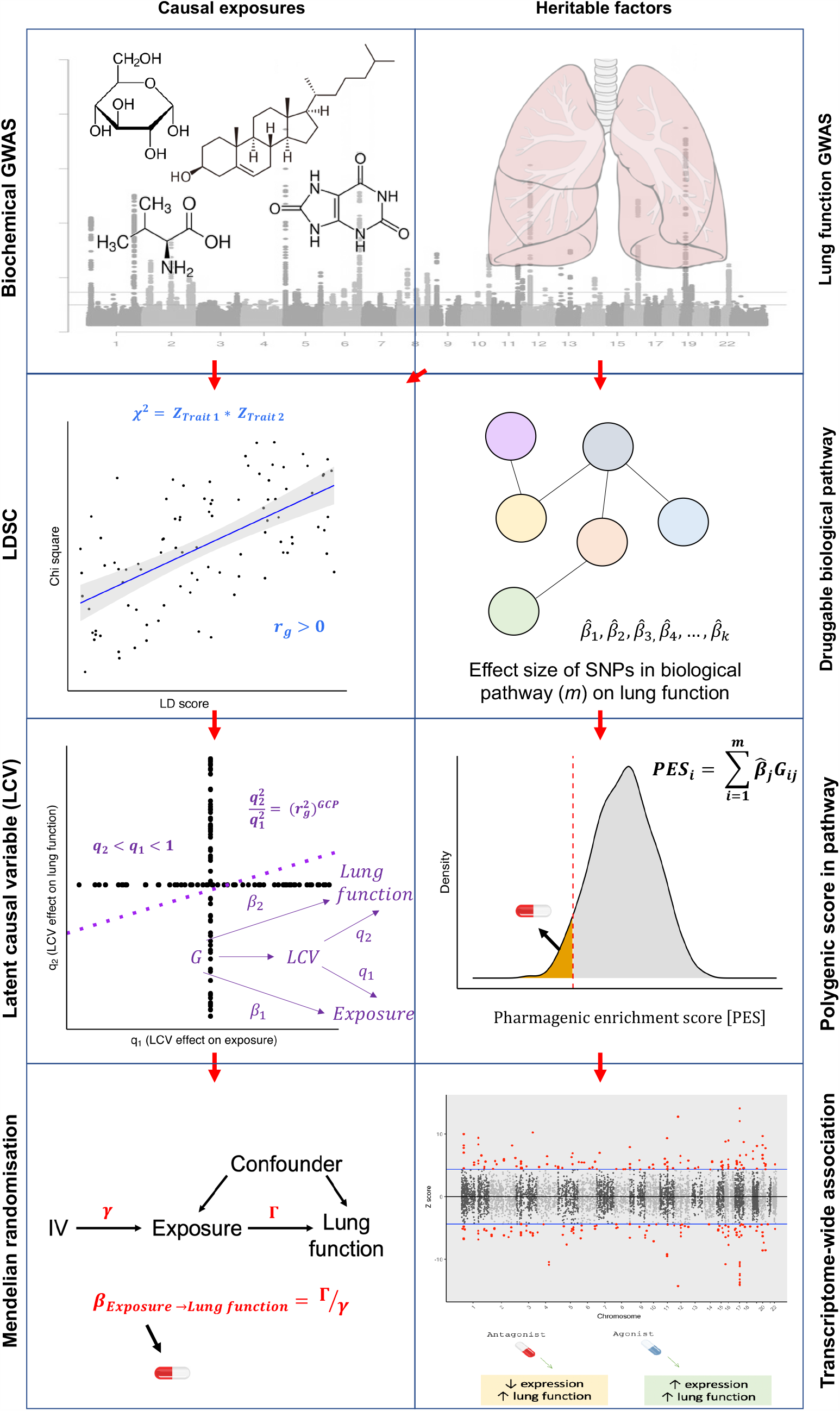
Overview of strategies for genetically informed drug repurposing to improve lung function. The left column outlines our workflow for using causal inference to identify drug targets, while the right side shows the workflow for functionally partitioning the heritable component into drug targets. In both cases we utilize or integrate GWAS data for lung function (including three spirometry phenotypes: forced expiratory volume in one second (FEV_1_), forced vital capacity (FVC), and their ratio (FEV_1_/FVC)) and quantitative biochemical traits (e.g. hormones and metabolites) which can be pharmacologically modulated. Using this data, we established genetic correlation between lung function and the biochemical traits using LD score regression (LDSC) (left column). We then constructed a latent causal variable (LCV) model to investigate evidence of causality for significantly correlated biochemical-lung function trait pairs. To further support causal inference between significant pairs we implemented Mendelian randomisation. Where a causal relationship between a modifiable biochemical trait and lung function is established, we can infer a novel treatment. The right column shows the workflow for utilising the *pharmagenic enrichment score* (PES) framework for precision drug repositioning. Specifically, polygenic scores for lung function were calculated using lung function GWAS SNPs within biological pathways that can be targeted by approved drugs, rather than a genome-wide score. Individuals with low genetically predicted lung function by a PES (low PES) relative to a reference population (orange shaded distribution in right panel 3) may benefit from a compound which modulates said pathway. To further support putative genetically predicted targets for drug repositioning a transcriptome-wide association study (TWAS) of lung function was performed. Druggable genes for which genetically predicted expression was correlated with a spirometry measure. Genes with positive genetic covariance between imputed expression and lung function (i.e. increased expression associated with increased lung function) could be modulated by an agonist compound, whilst genes for which decreased predicted expression is associated with improved lung function could be targeted by an antagonist compound.

## RESULTS

### Measures of lung function were genetically correlated with clinically significant metabolites and hormones

We assessed genetic correlation between three pulmonary function measurements (FEV_1_, FVC, and FEV_1_/FVC) and 172 GWAS summary statistics of European ancestry using bivariate linkage disequilibrium score regression (LDSC)^13,14^. A number of clinically significant traits displayed significant genetic correlation with FEV_1_, FVC, and/or FEV_1_/FVC after the correcting for the number of tests performed (*P* < 2.9 × 10^−4^, Figure 2a, Supplementary Tables 1-3). FVC had the largest number of genetic correlations which surpassed Bonferroni correction (N = 35), followed by FEV_1_ and FEV_1_/FVC for which 25 and 8 traits survived multiple testing correction, respectively. The trait most significantly correlated with both FEV_1_ and FVC was waist circumference - FEV_1_: *r*_*g*_ = −0.19, *SE* = 0.02, *P* = 5.71 × 10^−20^, FVC: *r*_*g*_ = - 0.24, *SE* = 0.02, *P* = 9.54 × 10^−33^. Asthma demonstrated the most significant correlation with FEV_1_/FVC (*r*_*g*_ = −0.35, *SE* = 0.05, *P* = 3.49 × 10^−12^), which is expected given its significant negative correlation with FEV_1_ (*r*_*g*_ = −0.34, *SE* = 0.06, *P* = 7.43 × 10^−10^) but its relationship with FVC did not survive Bonferroni correction (*r*_*g*_ = −0.18, *SE* = 0.05, *P* = 1 × 10^−3^).

**Figure 2.**
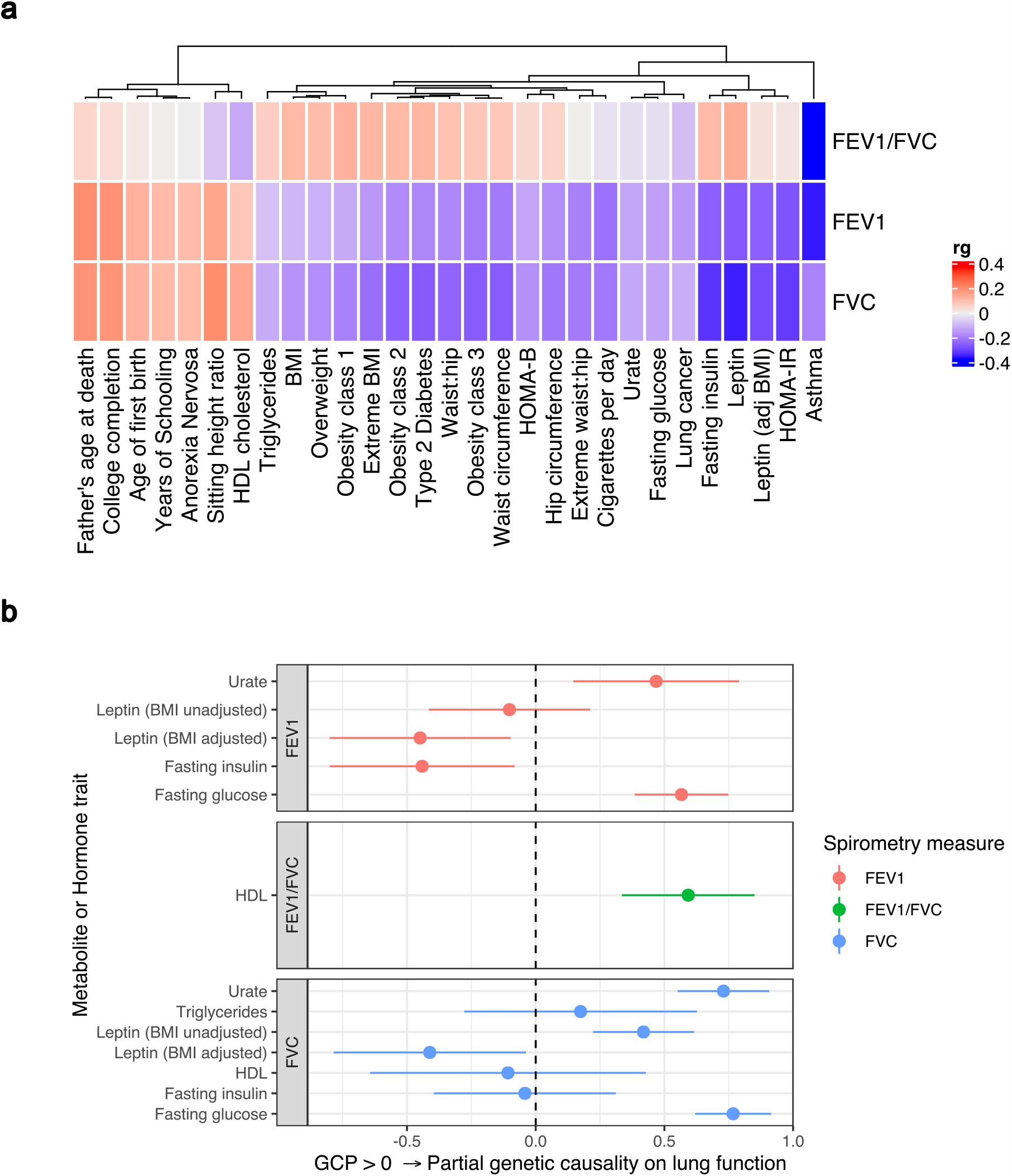
Genome wide investigation of biochemical traits related to lung function. (a) Heatmap of genetic correlations (r_g_) between three spirometry measures (FEV_1_, FVC, and FEV_1_/FVC) and a number of European ancestry GWAS. Genetic correlation estimates were plotted if the trait was significantly correlated with at least one of the lung function traits after Bonferroni correction. Hierarchical clustering was applied to the rows and utilised Pearson’s correlation distance. (**b**) Latent causal variable models between correlated biochemical traits (selected by LD score regression) that are potentially drug targets (metabolite or hormone traits) and each measure of lung function. The mean posterior genetic causality proportion (GCP) is plotted, with the error bars representing the upper and lower limits defined by its standard error. A positive GCP estimate significantly different than zero indicates partial genetic causality of the biochemical trait on the spirometry measure.

Interestingly, there was evidence of genetic correlation between measures of lung function and circulating levels of both metabolites and hormones. This is notable as these molecules can be pharmacologically modulated, potentially informing novel therapeutic strategies and drug repurposing opportunities to improve lung function. Significant genetic correlations were observed with four metabolites (fasting glucose, high-density lipoprotein [HDL], triglycerides, and urate) and two hormones (fasting insulin and leptin) for at least one measure of lung function (Table 1). These significant relationships were as follows: FEV_1_ was negatively correlated with fasting insulin, leptin (adjusted and unadjusted for BMI), urate, and fasting glucose; FVC was negatively correlated with the same traits as FEV_1_ but was further positively correlated with HDL and negatively correlated with circulating triglycerides; FEV_1_/FVC was conversely negatively correlated with HDL.

**Table 1:** **Significant genetic correlations between lung function measures and metabolite and hormone GWAS**

| Lung function trait* | Biochemical trait | Genetic correlation ($r_g$)<sup>#</sup> |
| --- | --- | --- |
| FEV<sub>1</sub> | Fasting insulin | -0.23 (0.04) |
| FEV<sub>1</sub> | Leptin (BMI unadjusted) | -0.25 (0.05) |
| FEV<sub>1</sub> | Leptin (BMI adjusted) | -0.24 (0.05) |
| FEV<sub>1</sub> | Urate | -0.12 (0.03) |
| FEV<sub>1</sub> | Fasting glucose | -0.13 (0.03) |
| FVC | Fasting insulin | -0.31 (0.04) |
| FVC | Leptin (BMI unadjusted) | -0.33 (0.05) |
| FVC | Leptin (BMI adjusted) | -0.27 (0.05) |
| FVC | HDL cholesterol | 0.14 (0.03) |
| FVC | Urate | -0.12 (0.02) |
| FVC | Triglycerides | -0.11 (0.03) |
| FVC | Fasting glucose | -0.12 (0.03) |
| FEV<sub>1</sub>/FVC | HDL cholesterol | -0.11 (0.03) |

\*FEV<sub>1</sub> = forced expiratory volume in one second, FVC = forced vital capacity, FEV<sub>1</sub>/FVC = ratio of FEV<sub>1</sub> to FVC. <sup>#</sup>Genetic correlations which survived multiple testing correction for each lung function trait individually are reported with their respective standard error.

### Evidence of a causal relationship between fasting glucose and lung function supports antihyperglycaemic compounds as drug repurposing candidates

The genetic correlations observed between lung function measures and metabolite/hormone traits may be clinically actionable, however, a significant estimate of genetic correlation does not imply causality^15^. In response, we constructed a latent causal variable (LCV) model to estimate mean posterior genetic causality proportion 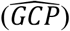 for each metabolite or hormone trait and the lung function measure with which it is genetically correlated (Figure 2b, Supplementary Table 4). The LCV method assumes that a latent variable mediates the genetic correlation between two traits, and tests whether this latent variable displays stronger correlation with either of the traits. Specifically, the mixed fourth moments of the bivariate effect size distributions for all SNPs and their LD structure are leveraged to derive a mean posterior estimate of the GCP, which quantifies the magnitude of genetic causality between the two traits. GCP values range from −1 to 1 (full genetic causality), within these limits positive values indicate greater partial genetic causality of trait one on two, and vice versa for negative values. We tested whether the mean posterior GCP estimate was significantly different from zero as evidence of partial genetic causality. Firstly, considering fasting glucose and both FEV_1_ and FVC the mean posterior GCP estimate was significantly different from zero, and positive, which suggested a partial causal effect of fasting glucose on these two measures of lung function, with the causal relationship more rigorous with 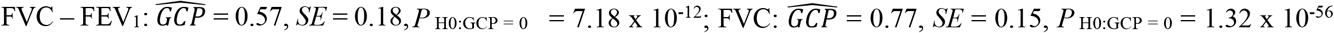. Posterior mean GCP estimates were marginally attenuated but remained significant upon using BMI adjusted fasting glucose estimates 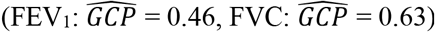. In addition, partial genetic causation was detected between HDL cholesterol and FEV_1_/FVC 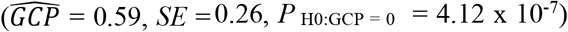 and leptin with 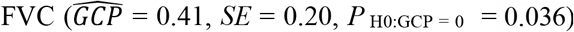, although these models displayed comparatively weaker evidence. A non-zero mean posterior GCP estimate was observed for urate and 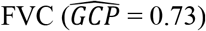, however, the relatively low heritability *z* score as calculated by the LCV framework (*z* < 7) may lead to an inflated estimate. There was no significant evidence of genetic causality between any of the remaining LDSC prioritised hormone or metabolite traits and FEV_1_, FVC, or FEV_1_/FVC.

As it was the most significant LCV model, the causal effect of fasting glucose on FEV_1_ and FVC was further investigated utilising a Mendelian randomisation (MR) approach. MR differs from an LCV model as it exploits genome-wide significant variants as genetic instrumental variables (IV) to calculate a causal estimate of an exposure (fasting glucose) on an outcome (lung function). We selected 32 genome-wide significant variants associated with glucose in approximate linkage equilibrium as IVs (*P* < 5 × 10^−8^, *r*^2^ < 0.001) to ensure that variants were both rigorously associated with the exposure and independent from one another. A 1 mmol/L increase in fasting glucose was associated with a −0.088 (95% CI: −0.17, −0.01) standard deviation decline in FVC using an inverse variance weighted (IVW) estimator with multiplicative random effects. Similarly, elevated fasting glucose was also shown to have a negative effect on FEV_1_: *β*_IVW_ = −0.096 [95% CI: −0.18, −0.01]. The causal estimate for fasting glucose was nominally significant for both FVC and FEV_1_ (*P* = 0.033 and *P* = 0.023, respectively) using an IVW estimator with multiplicative random effects. It should be noted that implementing the IVW model with fixed effects yields narrower confidence intervals for the respective FVC and FEV_1_ beta coefficients: FVC – *β*_IVW_ = −0.088 [95% CI: −0.12, −0.05], *P* = 2.18x 10^−6^; FEV_1_ – *β*_IVW_ = −0.096 [95% CI: −0.13, −0.06], *P* = 1.78 × 10^−7^ – however, there was evidence of statistical heterogeneity amongst the IV effects, indicating that the multiplicative random effects estimator was more appropriate (Cochran’s *Q*: *P* < 0.05)^3,16^. We implemented a number of sensitivity analyses to test the rigour of our causal estimate of the effect of fasting glucose on lung function (Figure 3, Supplementary Tables 5-7). Firstly, we obtained an analogous, and statistically significant, causal estimate using the weighted median method (FVC: *β*_Weighted median_ = −0.09 [95% CI: −0.16, −0.04], FEV_1_: *β*_Weighted median_ = −0.07 [95% CI: −0.13, −0.01]). The weighted median method relaxes the assumption that all IVs must be valid, as described elsewhere^17^. An MR-Egger model was then constructed, which includes a non-zero intercept term which can be used as a measure of unbalanced pleiotropy^18^. The causal estimate using MR-Egger was in the same direction for FEV_1_ and FVC, however, was non-significant (Supplementary Table 6). It should be noted that the MR-Egger method has notably less power than the IVW approach, particularly when fewer instruments are used^18^. Importantly, the MR-Egger intercept was not significantly different from zero in the FEV_1_ or FVC model, indicating no evidence of unbalanced pleiotropy. This was supported by a non-significant global test of pleiotropy implemented as part of the MR PRESSO (MR-Pleiotropy Residual Sum and Outlier) framework (Supplementary Table 6)^19^.

**Figure 3.**
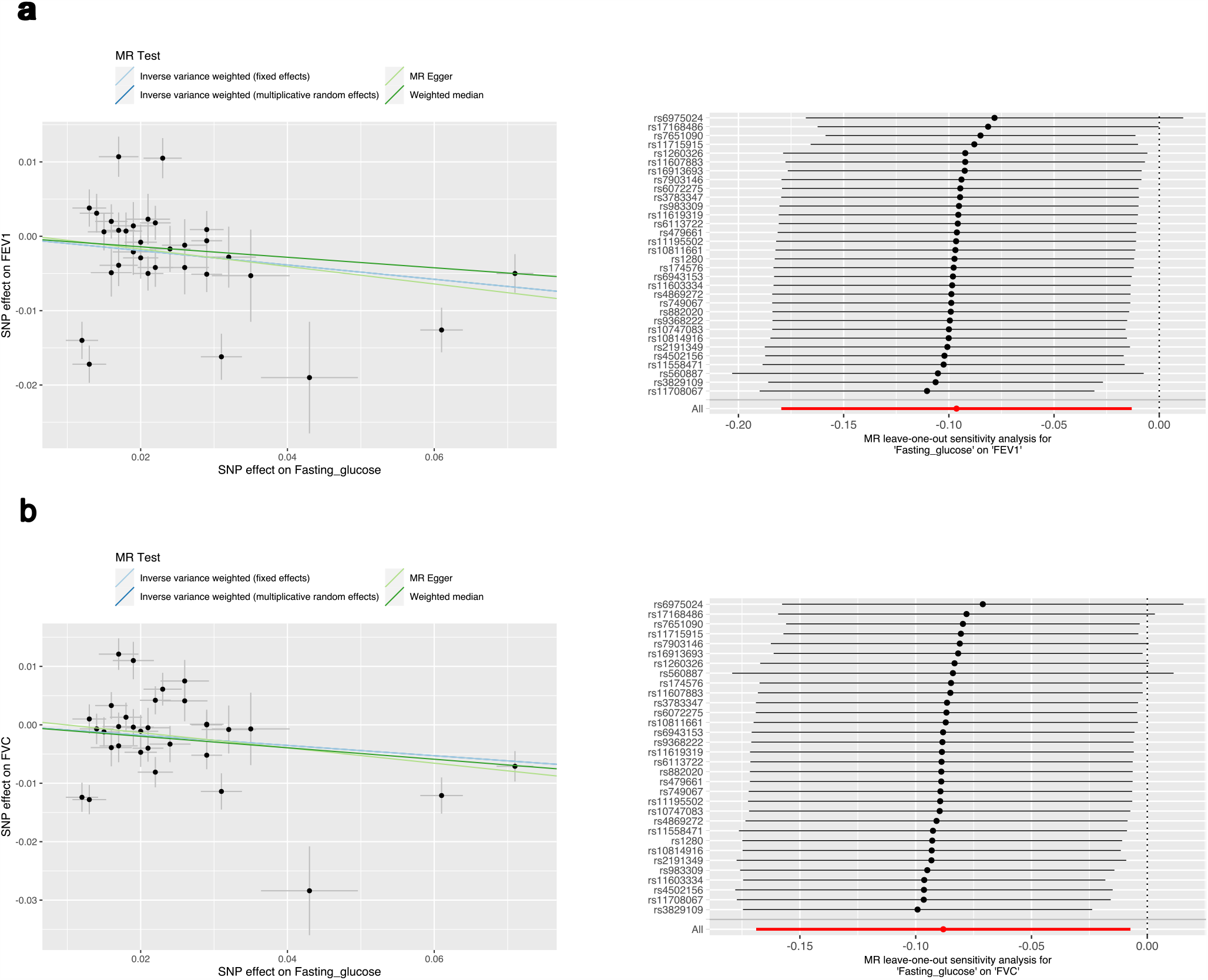
Investigation of the effect of fasting glucose on lung function using two-sample Mendelian randomisation. Mendelian randomisation (MR) results for (**a**) FEV_1_ and (**b**) FVC. The left scatter plot indicates the effect size of the instrumental variable (IV) SNP effects on fasting glucose (mmol/L) and FEV_1_ or FVC, respectively (standard deviation units). Each regression slope corresponds to the causal estimate between fasting glucose and each lung function measure, with the slopes shaded by the MR method used to calculate the causal estimate. The forest plot on the right indicates the results of the ‘leave one out analysis’. Each black point represents the causal estimate (IVW estimator with multiplicative random effects) of fasting glucose on each lung function measure with the SNP (IV) labelled on the y-axis removed; error bars represent upper and lower confidence intervals. The combined IVW estimate with all IVs is represented by the red point (‘All’).

Finally, we successively recalculated the IVW causal estimate for the effect of fasting glucose on FEV_1_ and FVC by removing one IV at a time in a ‘leave-one-out’ analysis (Figure 3, Supplementary Table 7)^20^. An analogous causal estimate was derived regardless of which IV was removed, however, there were five IVs (FEV_1_ model = two outlier SNPs, FVC model = four outlier SNPs, [two outlier SNPs shared]) for which the estimate was marginally non-significant after exclusion (maximum *P* = 0.11, IVW with multiplicative random-effects). We then used a phenome-wide association approach to demonstrate that these five SNPs were, i) annotated to genes with important roles in glycaemic homeostasis, and ii) were almost exclusively associated with glycaemic traits or diabetes (Supplementary Note, Supplementary Tables 8-12). As a result, we concluded that these IVs did not likely represent horizontal pleiotropy, which would bias the causal estimate, but instead were biologically salient IVs with large effects.

Whilst smoking status (ever vs never smoked) was a covariate in the lung function GWAS, we sought to assess whether the relationship between blood glucose and lung function could be driven by residual effects of smoking. There was a significant genetic correlation between the number of cigarettes smoked per day and fasting glucose (*r*_*g*_ = 0.16, *SE* = 0.043), although this was not observed with the ‘ever vs never smoked’ phenotype (*r*_*g*_ = 0.007, *SE* = 0.039). However, a latent causal variable model constructed for fasting glucose and cigarettes smoked per day did not indicate evidence of genetic causality, in contrast to the glucose/lung function models - 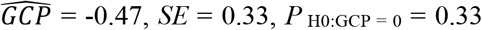. The MR IVs for glucose were further checked for association with either ‘ever vs never smoked’ and ‘cigarettes per day’, with none of the IVs demonstrating any association with either smoking phenotype at a genome-wide (*P* < 5 × 10^−8^) or suggestive (*P* < 1 × 10^−5^) significance threshold (Supplementary Tables 13,14). In summary, these data suggested there is an effect of fasting glucose on lung function beyond what is directly attributable to a residual impact of smoking.

### Implementation of the *pharmagenic enrichment score* for genetically informed drug repurposing in respiratory distress

We aimed to further expand drug repurposing opportunities for lung function using the *pharmagenic enrichment score* (PES) approach (Online methods)^12^. Briefly, PES aims to implement genetically informed drug repurposing with polygenic scores (PGS) calculated using genetic variants specifically within druggable pathways (Figure 4a). In the context of this study, individuals with a depleted PES for lung function (lower genetically predicted lung function) mapped to pathways with known drug targets may specifically benefit from drugs which modulate these pathways. The PES approach differs from a traditional genome wide PGS by providing direct biological insights into the potential impact of trait associated variation residing in drug function related gene-sets rather than the undifferentiated sum total of trait associated variation. Candidate pathways for the generation of PES profiles are obtained using GWAS summary statistics by identifying druggable gene-sets which display an enrichment of common variant associations. Firstly, we performed gene-set association of FEV_1_ and FVC using a collection of high-quality gene-sets from the molecular signatures database (MSigDB). These sets contain at least one gene which is modulated by an approved pharmacological agent (N_Sets_ = 1030, Online Methods). The FEV_1_/FVC phenotype is less directly interpretable in this context, given that it is used primarily as a diagnostic tool rather than as a quantitative measure, and thus, we focused on repurposing candidates for FEV_1_ and FVC individually. Previously, we extended the concept of *P*-value thresholding (*P*_T_) for PGS to the multi-marker gene level test-statistic and implemented this in our gene set analysis^12^. We argue that distinct biological processes in individuals may only be captured when the optimal spectrum of polygenic variation is included in the model. A variety of *P*_T_ could be utilised; for simplicity, we selected four *P*-values thresholds (all SNPs, *P*_T_ < 0.5, *P*_T_ < 0.05, and *P*_T_ < 0.005), in accordance with our previous work^12^. We annotated variants to genes using genomic proximity. Genic boundaries were extended to capture regulatory variation, with both conservative and liberal upstream and downstream boundary definitions. This involved an extension of 5 kilobases (kb) upstream of the gene, and 1.5 kb downstream for the conservative construct, whilst a larger 35 kb upstream and 10 kb downstream was implemented in the more liberal construct.

**Figure 4.**
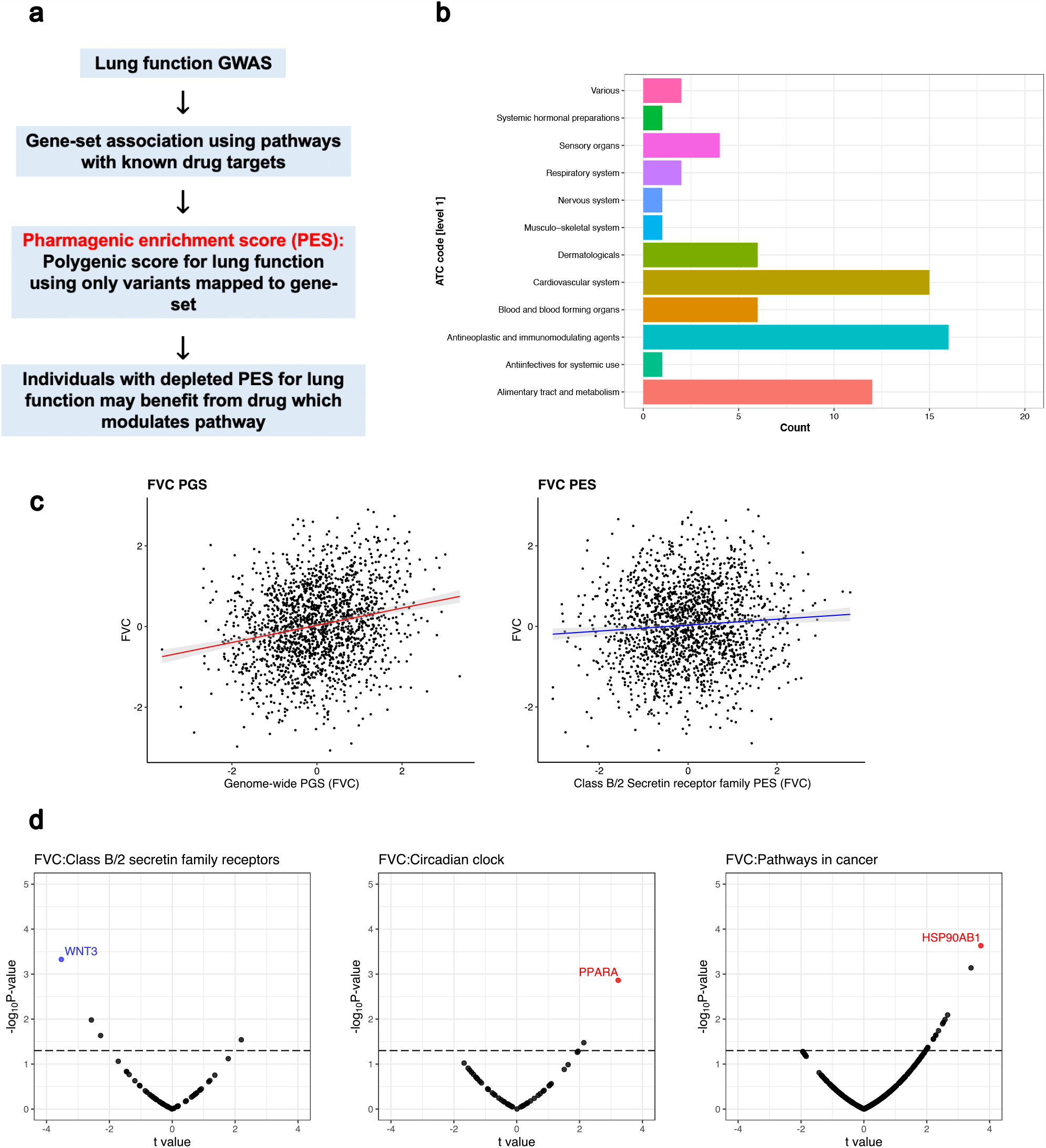
The *pharmagenic enrichment score* framework to identify and implement drug repurposing candidates for lung function. (**a**) Overview of the *pharmagenic enrichment score* (PES) approach, whereby polygenic scores of lung function measures are constructed using variants specifically within druggable pathways. Individuals with a depleted PES, that is, lower genetically predicted spirometry measures using variants in the gene-set, may benefit from a drug which modulates the pathway in question. (**b**) The number of FDA-approved drugs with overrepresented targets in at least one candidate PES gene-sets per anatomic therapeutic classification (ATC) level one code. Each ATC level one code is shaded a different colour with its frequency on the x-axis. (**c**) The phenotypic association between a polygenic score (PGS) of FVC and an FVC PES which was significant after correction for genome wide PGS. The relationship between the PES/PGS and normalised residual FVC in an independent cohort is plotted, with 95% confidence intervals of the regression trendline indicated by shading (**d**) Significant correlations between the expression of genes in a candidate PES and three lung function PES (FVC): *Class B/2 Secretin family receptors, Circadian clock*, and *Pathways in cancer*. The relationship between PES and gene-expression is presented as a volcano plot, where the x-axis is the *t* value (coefficient divided by standard error) and the y-axis is the -log_10_ *P*-value, with higher points more significant. Genes which are associated after multiple-testing correction for the number of genes in the pathway are coloured blue (strict FDR < 0.05) or red (lenient FDR < 0.1). The dotted line denotes an uncorrected nominally significant association (*P* < 0.05).

Gene-set association using the FEV_1_ and FVC GWAS was undertaken at each *P*_T_ with both conservative and liberal genic boundaries. If a gene-set was significant at multiple *P*_T_, the most significantly associated *P*_T_ was retained. The conservative genic-boundaries only yielded one druggable gene-set enriched with FEV_1_ associated variants after multiple testing correction (*q* < 0.05): *Signalling events mediated by the Hedgehog family* - *β* = 0.973, *SE* = 0.2, *P* = 9.3 × 10^−7^, *P*_*T*_ < 0.5, N_Genes_ = 22. There were no gene-sets with known drug targets using conservative genic-boundaries which survived multiple testing correction for association with FVC, however, several gene-sets trended towards significance (*q* < 0.1) including: *Pathways in cancer, TGF-beta signalling pathway*, and *signalling events mediated by the Hedgehog family* (Supplementary Table 15). Extending the genic boundaries to capture more regulatory variation (liberal boundaries) uncovered more druggable gene-sets (Supplementary Table 16). Specifically, there were seven and nine unique gene-sets which survived correction for FEV_1_ and FVC respectively (*q* < 0.05, Table 2). It should be noted that there were two pathways related to Hedgehog signalling, however, as these were from different annotation sources, and had a different number of genes, we considered them separately. A number of biological processes were encompassed by these prioritised gene-sets, such as: cancer (*Pathways in cancer, Basal cell carcinoma*), transforming growth factor (TGF)-beta superfamily signalling (*TGF-beta signalling pathway, BMP [Bone morphogenetic protein] receptor signalling, ALK [activin receptor-like kinase] in cardiac myocytes*), and cardiac function (*Dilated cardiomyopathy*).

**Table 2:**
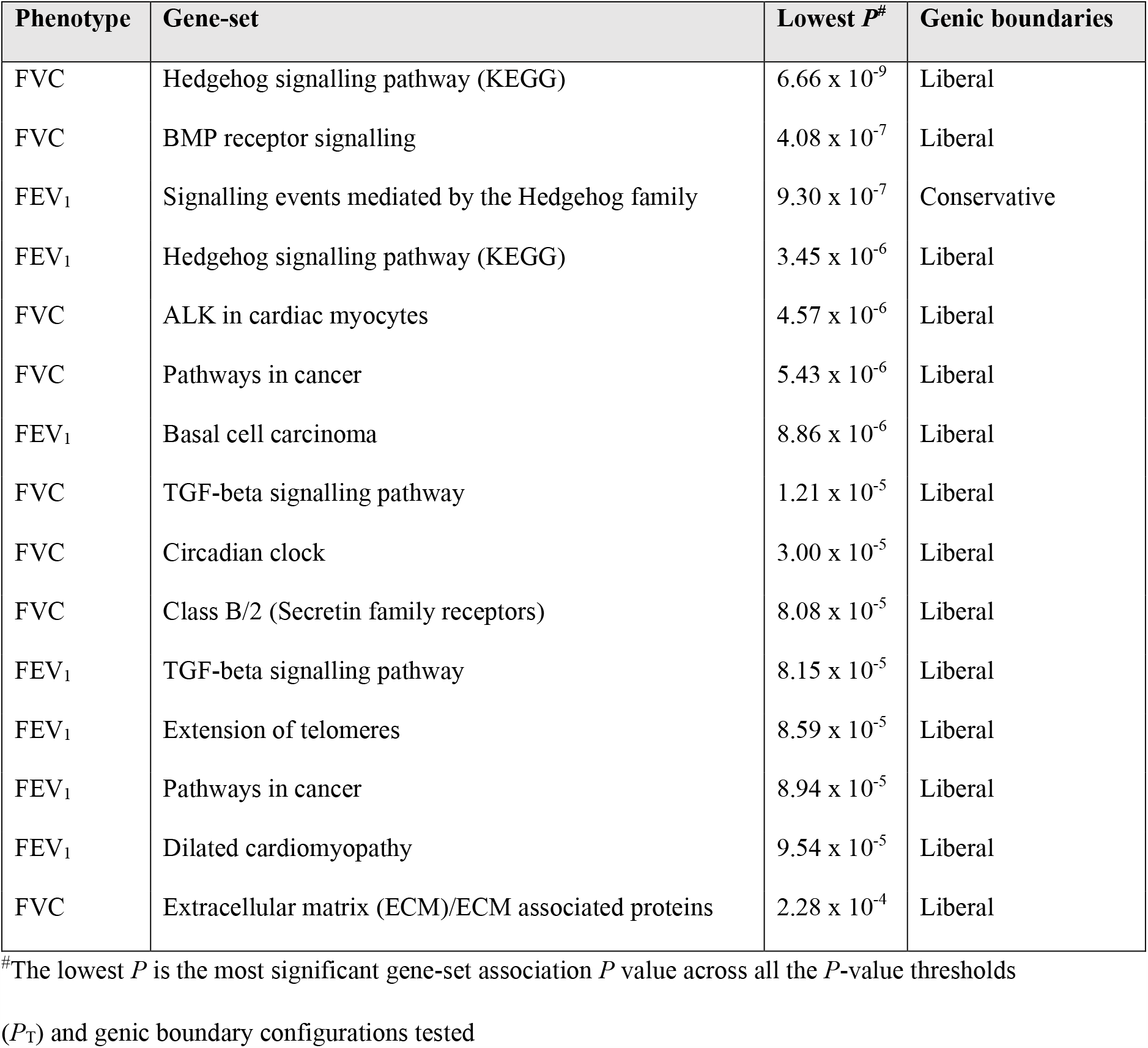
**Gene-sets with known drug targets enriched with lung function associated common variation after the application of multiple testing correction**

| Phenotype | Gene-set | Lowest $P^{\#}$ | Genic boundaries |
| --- | --- | --- | --- |
| FVC | Hedgehog signalling pathway (KEGG) | $6.66 \times 10^{-9}$ | Liberal |
| FVC | BMP receptor signalling | $4.08 \times 10^{-7}$ | Liberal |
| FEV<sub>1</sub> | Signalling events mediated by the Hedgehog family | $9.30 \times 10^{-7}$ | Conservative |
| FEV<sub>1</sub> | Hedgehog signalling pathway (KEGG) | $3.45 \times 10^{-6}$ | Liberal |
| FVC | ALK in cardiac myocytes | $4.57 \times 10^{-6}$ | Liberal |
| FVC | Pathways in cancer | $5.43 \times 10^{-6}$ | Liberal |
| FEV<sub>1</sub> | Basal cell carcinoma | $8.86 \times 10^{-6}$ | Liberal |
| FVC | TGF-beta signalling pathway | $1.21 \times 10^{-5}$ | Liberal |
| FVC | Circadian clock | $3.00 \times 10^{-5}$ | Liberal |
| FVC | Class B/2 (Secretin family receptors) | $8.08 \times 10^{-5}$ | Liberal |
| FEV<sub>1</sub> | TGF-beta signalling pathway | $8.15 \times 10^{-5}$ | Liberal |
| FEV<sub>1</sub> | Extension of telomeres | $8.59 \times 10^{-5}$ | Liberal |
| FEV<sub>1</sub> | Pathways in cancer | $8.94 \times 10^{-5}$ | Liberal |
| FEV<sub>1</sub> | Dilated cardiomyopathy | $9.54 \times 10^{-5}$ | Liberal |
| FVC | Extracellular matrix (ECM)/ECM associated proteins | $2.28 \times 10^{-4}$ | Liberal |

<sup>#</sup>The lowest $P$ is the most significant gene-set association $P$ value across all the $P$ -value thresholds
( $P_T$ ) and genic boundary configurations tested

For each candidate PES gene-set we performed computational drug selection to identify approved compounds predicted to modulate the enriched pathway. Firstly, we investigated FDA-approved pharmacological agents with a statistically significant overrepresentation of target genes in each of these sets (N_Overlap_ ≥ 3, *q* < 0.05, Online Methods). Drugs which target, i)n multiple gene-set members, and ii) more genes than expected by chance, were assumed to be particularly relevant for a biological pathway. There were six such gene-sets from the PES candidates which survived multiple testing correction enriched with the targets of an FDA approved compound (*Pathways in cancer, Dilated cardiomyopathy, Class B/2 [Secretin family receptors], Circadian clock, Extension of telomeres*, and *Extracellular matrix (ECM)/ECM associated proteins*, Supplementary Table 17) – notable drugs included: the anti-mineralocorticoid Spironolactone, antihyperglycaemic compounds (Rosiglitazone, Pramlintide), antihypertensives (e.g. Verapamil and Felodipine), antineoplastic agents (e.g. Bexarotene and Sunitinib), and nutraceuticals (Zinc, Vitamin E, and Doconexent). Each compound was annotated with its Anatomical Therapeutic Chemical (ATC) classification; the most common first level ATC code amongst these compounds was antineoplastic and immunomodulating agents (*L*, N = 16), followed by cardiovascular system (*C*, N =15), and alimentary tract and metabolism (*A*, N = 12; Figure 4b). Each of these compounds was subjected to expert curation by a pharmacist in relation to side-effects and prior literature evidence as detailed in Supplementary Table 18 (Online Methods, Supplementary Methods). Single drug-gene matching was undertaken for remaining PES candidate gene-sets lacking an approved compound with statistically overrepresented target, retaining drug-gene interactions with at least two lines of evidence from DGIdb (Supplementary Tables 19-30).

In order to test the phenotypic relevance of FEV_1_ and FVC PES profiles, we utilised an independent genotyped cohort from the Hunter Community Study (HCS, N = 1804, Online Methods). Firstly, we constructed a genome-wide PGS for FEV_1_ and FVC at six different *P*-value thresholds (Supplementary Table 31). The optimum FEV_1_ genetic score explained approximately 6.4% of the variance in FEV_1_ measured in the HCS cohort, whilst the FVC PGS explained approximately 5.7% of variance in FVC. Each of the seven PES profiles were tested for association with FEV_1_ and/or FVC both with and without adjustment for genome-wide PGS. Four of the PES considered had at least a nominally significant association with their respective spirometry measure (*P*_Empirical_ < 0.05 [10000 permutations], Table 3, Supplementary Table 32). The variance explained by the significant PES was between 0.4% - 0.7%, with the number of independent SNPs in these scores ranging from 76 to 16390. We then constructed a model which was adjusted for genome wide PGS at the same *P*_T_ as the PES and found that only the *Class B/2 secretin family receptor* FVC PES remained nominally significant (*β* = 0.047, *SE* = 0.022, *P* = 0.038, Figure 4c, Supplementary Table 33). This suggested that there was a relationship between the *Class B/2 secretin family receptor* FVC PES and FVC beyond what is attributable to a genome-wide PGS. This PES did not display any association with smoking status in this cohort (*β* = −0.014, *SE* = 0.047, *P* = 0.758). Furthermore, there was a significant depletion of FVC within the 10^th^ percentile (low genetically predicted FVC) of the *Class B/2 secretin receptor family* FVC PES in the HCS cohort, with the odds of being in the lowest decile decreasing by around 20% per standard deviation increase in FVC (OR = 0.80 [95% CI: 0.68, 0.93], *P* = 4.7 × 10^−3^). We further identified a subset of individuals with relative elevated FVC PGS (75^th^ percentile, higher genetically predicted FVC) but low phenotypic FVC (25^th^ percentile, normalised FVC residuals). The *Class B/2 secretin receptor family* FVC PES was assessed in these HCS participants (N = 71, 16.6% of 75^th^ percentile FVC PGS subset) to test whether there was a depletion of that spirometry measure, that is, low genetically predicted lung function using the PES. We found a non-significant trend of lower *Class B/2 secretin receptor family* PES amongst those with a high genome-wide burden of FVC increasing alleles but diminished FVC relative to the HCS cohort: *β* = −0.217, *SE* = 0.134, *P* = 0.106. All of the PES tested demonstrated small albeit significant correlations with genome wide PGS at the same *P*_T_ in the HCS cohort, with the exception of the *Extracellular matrix* PES for which the correlation was relatively large (*r* = 0.33, Supplementary Figure 1). The higher correlation in this gene-set was probably due to the large number of genes involved (>1000). Interestingly, there was still a number of individuals with high genetically predicted lung function using a genome wide PGS (90^th^ percentile of HCS cohort) but low genetically predicted lung function using one of the PES (10^th^ percentile). Specifically, 12.17% and 12.05% of the HCS participants in the 90^th^ percentile PGS for FVC and FEV_1_ respectively had a depleted PES (10^th^ percentile, low predicted lung function by PES). Taken together, this suggests that pathway based polygenic scores provide distinct biological insights for some individuals with otherwise high genetic load of lung function increasing alleles.

**Table 3:**
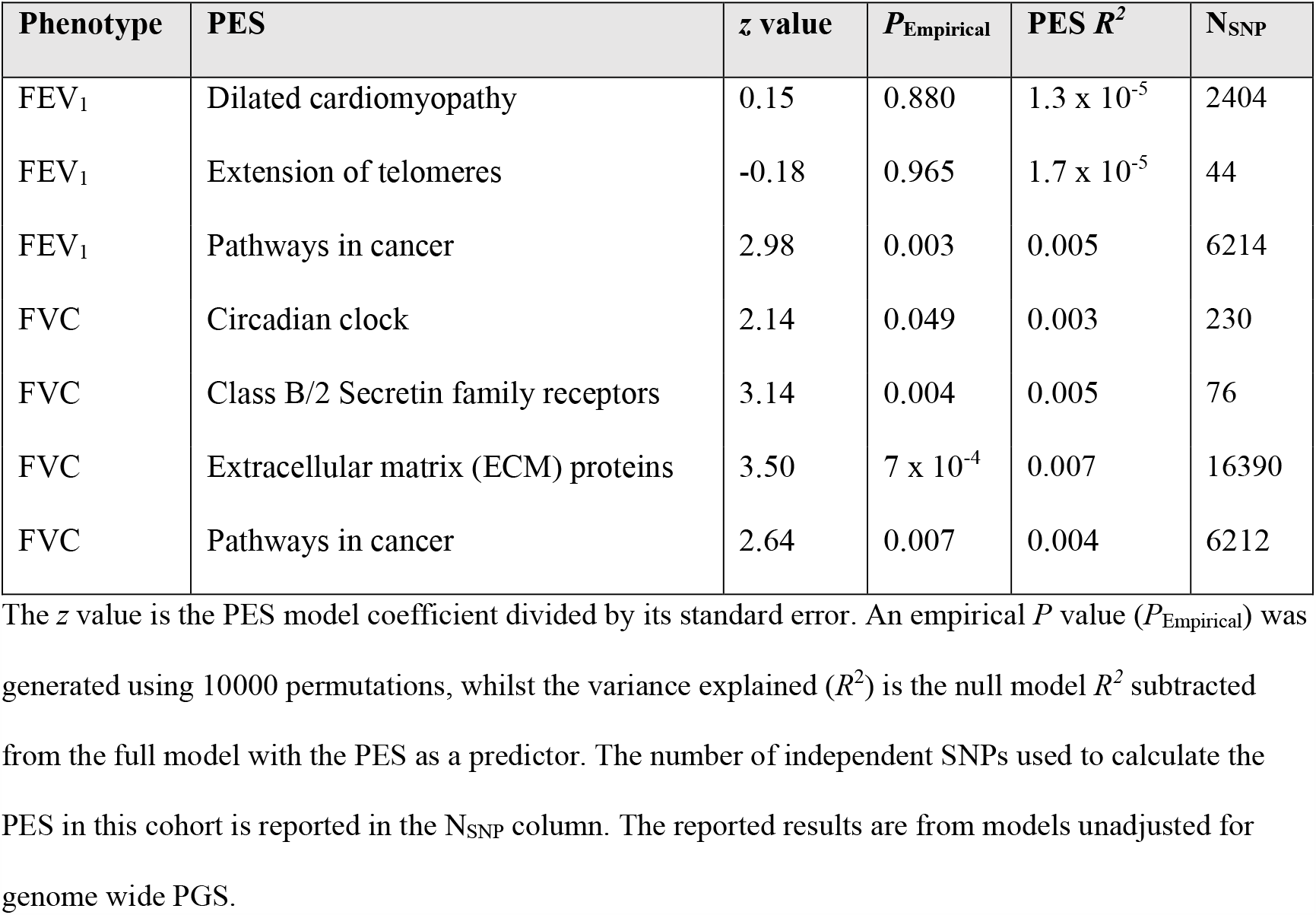
**The association between lung function PES and spirometry measures in the Hunter Community Study cohort**

| Phenotype | PES | *z* value | *P*<sub>Empirical</sub> | PES *R*<sup>2</sup> | N<sub>SNP</sub> |
| --- | --- | --- | --- | --- | --- |
| FEV<sub>1</sub> | Dilated cardiomyopathy | 0.15 | 0.880 | 1.3 x 10<sup>-5</sup> | 2404 |
| FEV<sub>1</sub> | Extension of telomeres | -0.18 | 0.965 | 1.7 x 10<sup>-5</sup> | 44 |
| FEV<sub>1</sub> | Pathways in cancer | 2.98 | 0.003 | 0.005 | 6214 |
| FVC | Circadian clock | 2.14 | 0.049 | 0.003 | 230 |
| FVC | Class B/2 Secretin family receptors | 3.14 | 0.004 | 0.005 | 76 |
| FVC | Extracellular matrix (ECM) proteins | 3.50 | 7 x 10<sup>-4</sup> | 0.007 | 16390 |
| FVC | Pathways in cancer | 2.64 | 0.007 | 0.004 | 6212 |

The *z* value is the PES model coefficient divided by its standard error. An empirical *P* value (*P*<sub>Empirical</sub>) was generated using 10000 permutations, whilst the variance explained (*R*<sup>2</sup>) is the null model *R*<sup>2</sup> subtracted from the full model with the PES as a predictor. The number of independent SNPs used to calculate the PES in this cohort is reported in the N<sub>SNP</sub> column. The reported results are from models unadjusted for genome wide PGS.

The correlation between the expression of genes within each pathway encompassed by the PES and the PES profiles themselves could provide further support for their biological impact. We investigated the association between lung function PES and gene expression using RNA sequencing (RNAseq) on transformed lymphoblastoid cell lines (LCL) from 357 European individuals for which phase 3 whole genome sequencing data was available from the 1000 genomes project (Online methods, Figure 4d, Supplementary Tables 34-40)^21^. We identified a significant association between the FVC PES *Class B/2 [Secretin family receptors]* and the expression of *WNT3* using a strict FDR threshold *q* < 0.05 (*t* = −3.53, *P* = 4.71 × 10^−4^, *q* = 0.028); a more lenient FDR cut-off (*q* < 0.1) yielded two more significant PES-gene expression correlations - FVC *Circadian clock* PES and *PPARA*: *t* = −3.23, *P* = 1.37 × 10^−3^, *q* = 0.07; FVC *Pathways in cancer* and *HSP90AB1*: *t* = 3.72, *P* = 2.33 × 10^−4^, *q* = 0.066. Expression of *WNT3* and *PPARA* were not associated with genome wide PGS at the same *P* value threshold (*P* = 0.63 and *P* = 0.29), whilst the PGS exhibited a weaker, nominal relationship with *HSP90AB1* (*P* = 0.04). The remaining four PES tested (FEV_1_ or FVC) all demonstrated at least one nominal, uncorrected association (*P* < 0.05). The observed effects of PES on gene expression at the population level were subtle; this is not surprising as each PES profile will encompass heterogenous variants for each individual, and thus, impacts on gene expression may be greater within specific genomic contexts.

### Transcriptome-wide association identifies putative targets for pharmacological modulation of lung function

We performed a transcriptome-wide association study (TWAS) of the three lung function measures using SNP weights from lung and blood tissue. TWAS leverages models of genetically regulated expression to test for a correlation between predicted expression and a phenotype^22^. Models of imputed expression derived from *cis*-eQTLs are generated from genes for which expression displays significant *cis*-heritability, that is, a significant genetic contribution to expression variance. We aimed to identify genes for which increased or decreased expression was associated with increased lung function and had approved compounds available which could improve lung function based on their mechanism of action (Figure 5a). For instance, if increased expression of a gene was associated with improved lung function, then an agonist of that gene may be clinically useful or vice versa in the case of decreased expression. Using a Bonferroni threshold for the number of genes tested in lung and blood individually, we identified a number of transcriptome-wide significant genes as follows - FEV_1_: N_Genes [Lung]_ = 232, N_Genes [Whole blood]_ = 201; FVC: N_Genes [Lung]_ = 222, N_Genes [Whole blood]_ = 167 (Supplementary Tables 41-44, Figure 5b). Transcriptome-wide associated genes were only retained if they were not also associated with a smoking phenotype, to minimise residual smoking related confounding. Specifically, we tested whether predicted expression of the genes which survived correction in the FEV_1_ or FVC TWAS were associated with smoking behaviour (‘ever vs never smoked’ and ‘cigarettes per day’) in a TWAS using SNP weights from lung, blood, and two brain regions implicated in nicotine addiction (dorsolateral prefrontal cortex and nucleus accumbens – Online Methods, Supplementary Tables 45-52)^23,24^. We searched each of these significant genes in the Drug-Gene Interaction Database (DGIdb v3.0.2) to ascertain compounds which may improve lung function based on the direction of effect from the TWAS analyses. In accordance with the PES analyses FEV_1_/FVC was not directly considered and we focused on FEV_1_ and/or FVC associated genes which could be pharmacologically modulated. A tiered system was utilised to select drug-gene interactions which may enhance lung function, whereby tier one were FDA approved compounds, and tier two were investigational (Online Methods, Supplementary Table 53).

**Figure 5.**
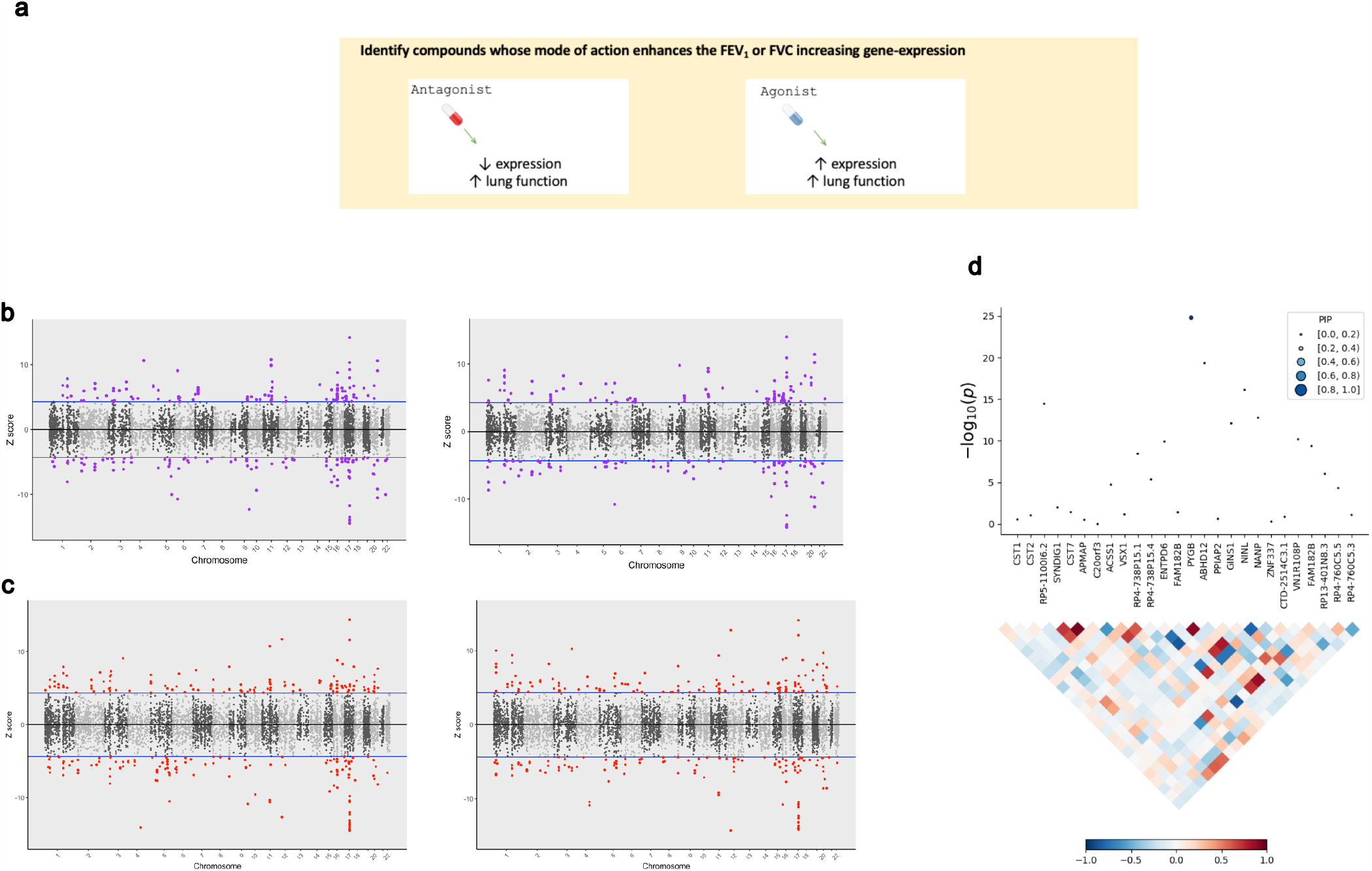
The application of transcriptome-wide association to identify drug repurposing candidates for lung function. (**a**) Schematic outlining the use of TWAS to reveal clinically actionable drug-gene interactions. Druggable genes with lung function associated imputed expression can be finemapped to prioritise a credible set of a causal genes at the TWAS locus, that is a high posterior inclusion probability (*PIP*). We seek to identify drugs with a mode of action which match the TWAS *Z* value, that is, compounds which may increase lung function. (**b** – **c**) Miami plots of a TWAS of FEV_1_ (left) and FVC (right) using whole blood (**b**) and lung (**c**) SNP weights. TWAS *Z* > 0 denotes a gene for which increased predicted expression is associated with increased lung function and vice versa. The highlighted genes survived multiple testing correction for the number of genes tested. (**d**) Probabilistic finemapping of the *PYGB* TWAS locus. The points denoting each gene are sized and coloured by their posterior inclusion probability for causality (PIP), with higher PIP denoted by larger, darker points as represented on the scale. The correlation plot below each region represents the covariance of predicted expression between genes.

Four candidate genes were identified satisfying tier one criteria: *PPARD, ADORA2B, KCNJ1*, and *AMT*. For instance, decreased expression of potassium channel gene *KCNJ1* was associated with FVC (*Z*_TWAS_ = −4.60), and this channel can be inhibited by approved compounds such as the antidiabetic drug glimepiride. There were an additional seven genes with tier two investigational targets: *PYGB, PIK3C2B, LINGO1, APH1A, OPRL1, MST1R*, and *ACVR2B*. Probabilistic finemapping of these transcriptome-wide significant regions using a multi-tissue reference panel was then performed to prioritise whether these genes are likely causal at that locus (Online Methods). A credible set with 90% probability of containing the causal gene was computed for each locus utilising the marginal posterior inclusion probability (*PIP*) calculated from the observed TWAS statistics. We did not proceed with finemapping the *PPARD* locus due to its proximity to the defined boundaries of the MHC region. Two FEV_1_ associated genes with tier one and/or tier two drug interactions, *AMT* and *PYGB*, were included in the credible set with a *PIP* > 0.9. Tetrahydrofolate is a co-factor for *AMT* (*Z*_TWAS_ = 5.96, *PIP*= 0.893, whole blood SNP weights), which has been previously implicated as having a beneficial effect on lung function. *PYGB* (*Z*_TWAS_ = −6.98, *PIP* = 0.999, lung SNP weights) encodes a protein involved in glycogenolysis and can be putatively inhibited by the new exploratory treatment for respiratory failure, Sivelestat (Figure 5c).

In addition, we tested a more conservative Bernoulli prior for each causal indicator (p = 1 × 10^−5^) but this only had a negligible effect on the posterior inclusion probability for either *AMT* (*PIP* = 0.87) or *PYGB* (*PIP* = 0.994). Whilst there is a plausible role for *AMT* in respiratory biology (Aminomethyltransferase, involved in glycine cleavage), it should be noted that decreased predicted expression of *AMT* also trended towards the Bonferroni threshold for a significant association with smoking status (*Z*_TWAS_ = −4.33, *P* = 1.46 × 10^−5^), although this was weaker for the cigarettes per day phenotype (*Z*_TWAS_ = −2.97, *P* = 2.94 × 10^−3^). As a result, the association of this region with FEV_1_ should be treated cautiously until its biological relevance can be clarified to ensure that this signal is not driven by a residual effect of smoking.

### Host-viral interactomes suggested proposed pulmonary drug repurposing candidates may be significant for respiratory virus infection

Respiratory viruses are an important contributor to acute, and potentially fatal, declines in lung function. We sought to investigate whether our proposed drug repurposing candidates for lung function may also exhibit anti-viral properties against these pathogens. The host-virus interactome was analysed for three respiratory viruses to perform computational drug repurposing – severe acute respiratory syndrome coronavirus 2 (SARS-CoV2), influenza (H1N1), and the human adenovirus (HAdV) family (Online methods, Supplementary Tables 54-56)^25–27^. Specifically, human proteins which are predicted to interact with virally expressed proteins (‘prey proteins’) were investigated to identify those which could be inhibited by existing drugs to potentially disrupt the progression of infection. Approved inhibitors or antagonists of proteins in each respective host-virus interactome were sourced using DGidb and compared to our candidate compounds for lung function from the PES approach. Furthermore, we investigated the reported drug-label side-effect frequencies of each of these overlapping pharmacological agents and retained only candidates with no commonly reported (> 1% frequency) respiratory adverse effects. There were three inhibitors of human proteins with evidence of interaction with a viral protein that also targeted a gene which was a member of a PES candidate gene-set. Vorinostat (*HDAC2* inhibitor) and Aminocaproic acid (*PLAT* inhibitor) both inhibited a SARS-CoV2 ‘prey protein’ and targeted a gene within the *Pathways in cancer* and *Extracellular matrix (ECM)/ECM associated proteins* PES pathways, respectively. Similarly, Ruxolitinib inhibits the influenza prey protein *JAK1*, a part of the *Pathways in cancer* gene-set.

We demonstrated using multiple lines of evidence a putative relationship between increased fasting blood glucose and lung function – therefore, we investigated whether any of the host-viral interactome members were enriched within biological pathways involved in glycaemic homeostasis. Interestingly, there was an overrepresentation of SARS-CoV2 ‘prey proteins’ amongst four gene-sets related to glucose metabolism, along with insulin and glucagon signalling pathways (Table 4). Fourteen SARS-CoV2 ‘prey proteins’ were members of at least one of these gene-sets, with a greater number of interactions amongst these genes than expected by chance (*P* = 4.42 × 10^−12^, Supplementary Table 57). We outline evidence for the potential role of these viral prey genes in glycaemic homeostasis in supplementary table 57. These data support emerging evidence that SARS-CoV2 infected patients with hyperglycaemia are at higher risk of morbidity and mortality^28^.

**Table 4:** **Overrepresentation of proteins which interact with viral SARS-CoV2 expressed proteins within glycaemic related pathways**

| Glycaemic gene-set | P-value |
| --- | --- |
| Glucagon-like Peptide-1 (GLP1) regulates insulin secretion | $7.02 \times 10^{-4}$ |
| Glucagon signalling in metabolic regulation | $2.33 \times 10^{-4}$ |
| Glucose metabolism | $2.69 \times 10^{-5}$ |
| Regulation of insulin secretion | $2.13 \times 10^{-3}$ |

None of the glycaemic ‘prey proteins’ were direct target of antidiabetic compounds, however, 57% of these proteins had a high confidence protein-protein interaction with antidiabetic target gene (Supplementary Table 58). For instance, *GNB1* putatively binds with a SARS-CoV2 non-structural proteins (Nsp7) that forms the part of the replicase / transcriptase complex, whilst this protein also demonstrated evidence of interacting with 15 proteins modulated by an antidiabetic compound – such as *GLP1R*, which is the primary target of GLP-1 analogues, including exenatide. Pharmacological interventions which seek to control blood glucose may have positive implications both in terms of improving baseline lung function and reducing the risk of adverse consequences after SARS-CoV2 exposure.

## DISCUSSION

This study demonstrated a variety of methods for which genomic data could be utilised to propose drug repurposing candidates, ranging from approaches which exploit genome wide variant effects, to the identification of candidate clinically significant drug-gene interactions. Lung function is a particularly relevant phenotype to study in this context as its aetiology is influenced by a variety of complex biological factors and it is a significant contributor to global morbidity and mortality. We uncovered a number of putative pulmonary drug repositioning opportunities, with the role of glycaemic regulation in pulmonary function particularly interesting from a therapeutic perspective. Our study suggests a causal relationship between blood glucose and lung function using a genome-wide (LCV), and instrumental variable (MR) approach, whilst downregulation of the glycogen phosphorylase *PYGB* was also associated with FEV_1_ after probabilistic finemapping of TWAS loci. These data support previous literature suggesting that declines in pulmonary function are overrepresented amongst individuals with diabetes and correlates with poor glycaemic control^5,29–31^; a phenomenon which has also been reported in non-diabetics^32,33^. There are a number of pathophysiological mechanisms postulated to underlie this relationship, including fibrosis mediated by hyperglycaemia accelerated epithelial-to-mesenchymal transition^34^, and aberrant inflammatory responses to dysglycaemia^35,36^. Respiratory sequalae after infection may also be significantly affected by dysregulation of glycaemic control. Acute hyperglycaemia is associated with a significant increase in morbidity and mortality amongst non-diabetic community-acquired pneumonia (CAP) patients, which further supports its utility as a treatment target^37–40^. Notably, even patients with mild hyperglycemia [serum glucose 6-10.99 mmol/L] have a purported elevated risk of death at 90 days following CAP diagnosis^37^, whilst the association between type 2 diabetes and poor pneumonia outcomes appears to be driven by glycaemic control^40^. Inflammation is likely to be an important component of glycaemic influenced adverse effects, for instance, the intracellular carbohydrate *O*-Linked β-*N*-acetylglucosamine has been recently linked to influenza-associated cytokine storms^41^. Our findings supported the relevance of glycaemia to respiratory infection through demonstrating that proteins which putatively interact with the SARS-CoV2 virus were overrepresented in glycaemic pathways. Whilst the viral prey proteins we identified as members of glycaemic pathways were not the direct targets of antihyperglycaemic agents, some interact with these compounds, although biological saliency of these interactions warrants future investigation. The presence of a viral-prey protein interaction also does not necessarily support its essentiality in the viral life cycle and further data are needed to support this. Furthermore, the viral prey proteins overrepresented in the glycaemic pathways were mostly genes such as nucleoporins and cAMP-dependent protein kinases which have pleiotropic regulatory roles spanning a number of biological systems. These data taken together support the utility of managing blood glucose in the clinical improvement of respiratory outcomes.

Targeted drug application and repurposing is by its very nature confounded by biological heterogeneity amongst individuals. This is likely particularly true in the case of complex traits as their polygenic genetic architecture provides the substrate for each individual to display a unique profile of trait-associated variation. In the second stream of this study we stratified the polygenic architecture of lung function into a series of druggable pathways to provide a framework for pathway specific genetic scores we designate the *pharmagenic enrichment score* (PES). We suggest that leveraging inter-individual genetic heterogeneity in this way will improve the precision application of novel drug repurposing. A number of interesting drug repositioning candidates had overrepresented targets amongst the candidate PES gene-sets. For example, magnesium sulfate had enriched targets in the *Dilated cardiomyopathy* PES and has previously shown promise as a repurposing candidate to improve pulmonary function in asthma^42,43^. Using an independent cohort, several PES profiles tested explained a small, but significant, percentage of variance in FEV_1_ and/or FVC. The *Class B/2 secretin family receptors* score for FVC was particularly noteworthy given that it remained significant after an adjustment for genome wide PGS. Interestingly, this gene-set features a number of proteins involved with glycaemic homeostasis, including antidiabetic drug targets glucagon-like peptide receptor 1 (*GLP1R*) and amylin receptors (*RAMP1, RAMP2*, and *RAMP3*).While all of the PES demonstrated significant correlation with genome wide PGS, in the majority of cases it was small (*r* < 0.2), suggesting that most of these functionally relevant foci of genomic risk in lung function GWASs were relatively independent of the total PGS. Importantly, we still identified individuals with high genetically predicted lung function using a genome wide PGS but observed low predicted lung function with a pathway-specific PES. This was supported by the observed correlation between the PES and related mRNA expression which was distinct from a genome wide PGS. Collectively, these data are consistent with the hypothesis that important treatment-related biology can be captured at a pathway level for individuals with or at risk of respiratory illness.

Taken together our approach provides template for genetically informed precision drug repositioning to improve lung function. The clinical implementation in its most basic form would involve common variant genotyping using a commercial SNP array followed by imputation and lung function PES based stratification of treatment options. This would be combined with other biochemical exposure measures, such as fasting glucose, that are causal risk factors and have approved treatments. To illustrate the clinical implementation of our strategy, we generated a schematic representation of individual heterogeneity in biochemical and genetic components of risk in lung function and related them to candidates for precision drug repositioning (Figure 6). We envisage that our approach to variant and exposure risk stratification can be applied more broadly to identify and implement precision drug repositioning in range of complex traits.

**Figure 6.**
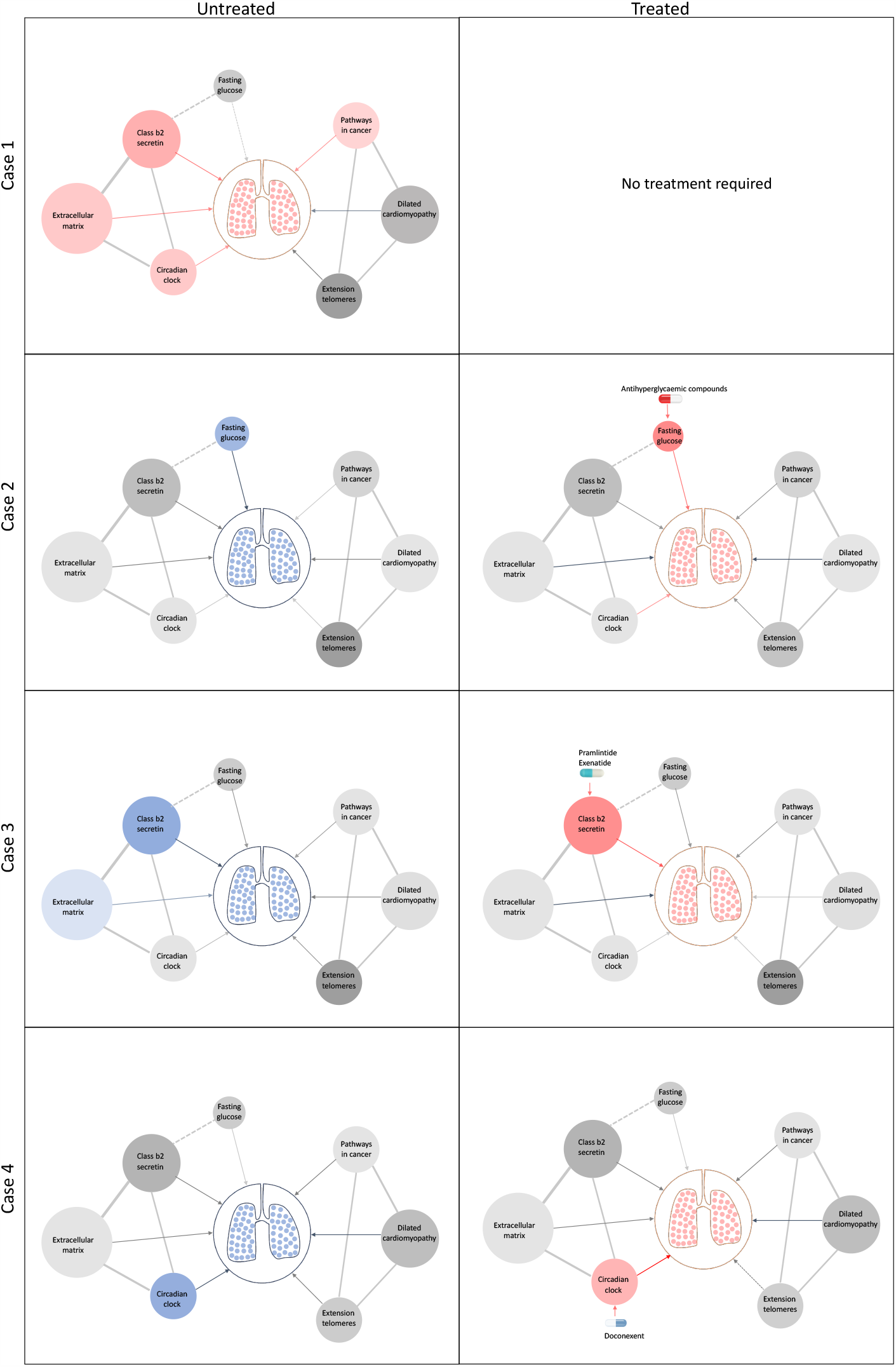
Schematic representation of drug repositioning and precision implementation in lung function deficits directed by causal enrichment of environmental and genetic risk factors. Each row represents a simulated individual with a heterogeneous presentation of risk factors related to lung function. Case 1 (top row) represents an individual with good lung function (pink lung tissue) and genomic and environmental components consistent with healthy lung function (grey to red nodes). These have a neutral to positive influence on lung function represented by the grey and red edges (arrow) respectively. Case 2 has high fasting glucose and neutral (grey) loading of genetic variants (PES) associated with lung function pathways. After treatment with antihyperglycaemic agents, or some other intervention to lower blood glucose, lung function is improved (red edge) sufficiently for therapeutic effect, represented by pink lungs. Case 3 has enrichment of genetic variants (PES) associated with poorer lung function in the *Class b2 secretin* pathway. To improve lung function, they are treated with drugs, such as pramlintide (which targets *RAMP1, RAMP2*, and *RAMP3*) and exenatide *(GLP1R* agonist), that works by modulating genes in the *Class b2 secretin* pathway to ameliorate the enrichment of poor lung function variants in that pathway. The broken edge between fasting glucose and the *Class b2 secretin* pathway represents the probable connection or shared genes between these nodes, as receptors in this pathway are involved in glycaemic regulation. Case 4 also presents with poor lung function (blue lung tissue) and enrichment of poor lung function associated variants in the *Circadian clock* pathway (blue node). This individual’s lung function was then treated by compounds, such as doconexent, that act on the *Circadian clock* pathway. This schematic is only representative of many thousands of treatment scenarios potentially informed by this treatment decision tool.

While there are some potential confounds in the use of GWAS data for causal inference via both latent causal variable models and Mendelian randomisation, such as, measurement error, population stratification, and horizontal pleiotropy, we are confident that the relationship between glycaemia and lung function presented in this study is robust given the multiple lines of support. Replicated, well-powered randomised controlled trials, however, are needed to fully resolve the clinical benefit of repurposing antihyperglycaemic compounds to improve lung function and in the context of viral infection. We also acknowledge that the direction of suitable pharmacological intervention is not inherently clear, such that an agonist or antagonist of genes within a pathway implicated by the PES approach is an important consideration^12^. Careful curation of proposed repurposing candidates will therefore be critical, particularly in the context of pulmonary traits where a variety of currently approved compounds have adverse respiratory effects. We suggest that TWAS could be utilised to help overcome these issues by identifying druggable genes which are members of candidate PES gene-sets for which a clinically beneficial impact on expression can be predicted. Interestingly, we also saw some evidence of cross talk between heritable risk at genes associated with lung function and fasting glucose, with the downregulation of the glycogen phosphorylase *PYGB* (associated with FEV_1_) observed through the probabilistic fine mapping of TWAS loci. In summary, we revealed candidate drug repurposing opportunities to potentially improve pulmonary function and provide the means for aligning their application in individuals that carry a high relative burden of variants associated with their function. Through this process we identify glycaemic interventions in particular, as being potentially beneficial in the context of respiratory infection.

## METHODS

### Lung function GWAS

We obtained GWAS summary statistics for FEV_1_, FVC, and their ratio from a meta-analysis of the UK biobank sample with the SpiroMeta consortium cohorts as outlined extensively elsewhere (N = 400102)^11^. Phenotypes were adjusted for age, age^2^, sex, height, smoking status (ever vs never smoked) and genotyping array before the residuals were subjected to rank inverse-normal transformation.

### Genetic correlation

Bivariate linkage disequilibrium score regression (LDSC) was performed between each lung function trait and a variety of GWAS as implemented by LDhub v1.9.3^14^. Lung function summary statistics were cleaned (‘munged’) prior to LDSC using munge_sumstats.py and merged with common HapMap3 SNPs excluding the MHC region due to its LD complexity, as is usual practice^13^. We retained estimates of genetic correlation (r_g_) for GWAS (N = 172) with European ancestry and a heritability *z* value > 4, as calculated by LDhub. When a phenotype had multiple GWAS, the GWAS with largest sample size was retained. The Bonferroni method was utilised for multiple testing correction - *P* < 2.9 × 10^−4^ (α = 0.05/172). A heatmap was constructed using the ComplexHeatmap package^44^.

### Latent causal variable models

Latent causal variable models were constructed between each measure of lung function which displayed a significant genetic correlation with a hormone or metabolite trait (see references for GWAS in supplementary table 4). The RunLCV.R and MomentFunctions.R scripts were leveraged to perform these analyses (https://github.com/lukejoconnor/LCV). The LCV framework assumes that a latent variable, *L*, mediates the genetic correlation between two traits (trait one, trait two), and uses the mixed fourth moments of the bivariate effect size distribution to estimate the mean posterior genetic causality proportion as described in detail by O’Connor and Price^15^. Specifically, the LCV model postulates that if trait two is partially causal for trait one, then directional SNP effects will be unequal, that is, variants impacting trait two will have a proportional effect on trait one, but this will not be observed in the other direction. The mean posterior GCP can be defined by equation one, where 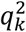 is the normalised effect of *L* on trait one or two respectively, and r_g_ is genetic correlation estimate:

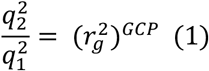

All traits were munged prior to LCV analyses, with only HapMap3 SNPs (MAF > 0.05) outside the MHC region retained in accordance with the LDSC analyses. We utilised the baseline 1000 genomes phase 3 LD scores for HapMap3 SNPs (MHC excluded). A two-sided *t* test was used to assess whether the estimated GCP was significantly different from zero.

### Mendelian randomisation

We investigated the causal effect of fasting glucose on both FEV_1_ and FVC using two-sample Mendelian randomisation (MR). MR is underpinned by the use of genetic variants as instrumental variables (IVs), with the random inheritance of these IVs as per Mendel’s laws facilitating the use of IVs to perform causal inference between an exposure and outcome, providing a series of assumptions are met^20^. These assumptions have been discussed previously^45^, briefly: the variant must be rigorously associated with the exposure; the variant must be independent of all confounders of the exposure-outcome relationship (“exclusion-restriction assumption”); and the variant must be associated with the outcome only by acting through the exposure. We satisfied the first assumption by selecting independent variants which are associated with fasting glucose using the traditional GWAS genome-wide significance threshold (*P* < 5 × 10^−8^, r^2^ < 0.001, palindromic SNPs removed). A different GWAS of fasting glucose was utilised for MR than for LDSC and LCV. Scott *et al*. performed a replication of ∼66000 Illumina CardioMetabochip variants following the Manning *et al*. GWAS for which more complete summary statistics were available, and thus, the later was included in the LDhub catalogue instead of the former^46,47^. We required only genome-wide significant SNPs for MR, therefore, the Scott *et al*. CardioMetabochip replication was more suitable as this was a larger sample size than the Manning *et al*. GWAS. Fasting glucose data for GWAS were obtained from either plasma or whole blood of non-diabetic individuals of European ancestry, and corrected to plasma levels (N = 133310, unit of effect = mmol/L)^47^. The remaining two IV assumptions cannot be definitively tested, and a suite of sensitivity analyses are implemented to provide evidence that they could be violated. Our primary MR model was an inverse-variance weighted effect model with multiplicative random effects^48^. Briefly, the pooled causal effect of the *j* IV exposure effects (*X*) on the outcome (*Y*) are estimated, where σ is the IV-outcome standard error (equation two).

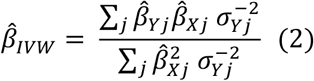

Further, we implemented a weighted median model which takes the median of the ratio estimates (as opposed to the mean in the IVW model), such that upweighting was applied to ratio estimates with greater precision^17^. An advantage of this approach is that it is subject to the ‘majority valid’ assumption, whereby an unbiased causal estimate will still be obtained if less than 50% of the model weighting arises from invalid IVs. An MR egger model was then constructed; an adaption of Egger regression wherein the exposure effect is regressed against the outcome with an intercept term (θ_0_) added to represent the average pleiotropic effect (equation three)^18^.

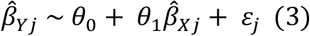

The key assumption of the MR egger model is referred to as Instrument Strength Independent of Direct Effect (InSIDE), which assumes that there is no significant correlation between direct IV effects on the outcome and genetic association of IVs with the exposure. In other words, the InSIDE assumption is violated if pleiotropic effects act through a confounder of the exposure-outcome association^16,18,49^. We also tested whether the Egger intercept is significantly different from zero as a measure of unbalanced pleiotropy or violation of the InSIDE assumption. In addition, heterogeneity amongst the IV ratio estimates was quantified using Cochran’s *Q* statistic, given that horizontal pleiotropy may be one explanation for significant heterogeneity. A global pleiotropy test was also implemented via the MR PRESSO framework^19^. Leave-one-out analyses were then performed to assess whether causal estimates are biased by a single IV, which may indicate the presence of outliers, and the sensitivity of the estimate to said outliers. However, outliers may not necessarily be evidence of horizontal pleiotropy. There were five IVs in either the FEV_1_ or FVC model where the IVW estimate was marginally no longer significant following their removal in the leave-one-out analysis. We performed a PheWAS for each of these SNPs using summary data collated by GWAS atlas v20191115 (https://atlas.ctglab.nl/) to assess evidence of horizontal pleiotropy, that is, acting through non-glycaemic pathways to influence lung function^50^. All MR analyses were performed in R version 3.6.0 using the TwoSampleMR v0.4.25 and MRPRESSO v1.0 packages.

### Investigating residual confounding from smoking on the relationship between fasting glucose and lung function

We investigated whether a residual effect of smoking could confound the link between glucose and lung function. Firstly, we selected two well-powered GWAS of smoking behaviours: ever vs never smoked (N = 385013)^50^, and cigarettes smoked per day (N = 263954)^51^. Genetic correlation between these two smoking phenotypes and fasting glucose was estimated as described above, followed by the construction of a latent causal variable model. The MR IVs utilised for fasting glucose were also checked for association with each smoking GWAS.

### Generation *pharmagenic enrichment score* (PES) candidate gene-sets

We implemented gene-set association using MAGMA method (MAGMA v1.06b), with some customisations to the framework to identify candidate PES genesets^12,52^. MAGMA aggregates SNP-wise *P* values for trait association into a gene-based *P* value and, thereafter, tests whether a set of genes is more strongly associated with the phenotype than all other genes. Gene-based test statistics were calculated analogous to Brown’s method, which is applicable to dependent *P*-values with known covariance (as common SNPs display through the phenomenon of linkage disequilibrium [LD], which can be quantified at a population level). Specifically, the mean χ^2^ gene test-statistic sums *P*-values mapped to each gene, using the 1000 genomes reference genotypes to scale the null χ^2^ distribution. *P*-value thresholding (*P*_T_) was utilised for the gene test statistic calculation; for simplicity, we selected four P-values thresholds (all SNPs, *P*_T_ < 0.5, *P*_T_ < 0.05, and *P*_T_ < 0.005). We mapped variants to 18297 autosomal genes in hg19 assembly defined by NCBI and obtained from the MAGMA website – genes within the major histocompatibility complex (MHC) were removed due to the complexity of LD within this region. The 1000 genomes phase 3 European reference panel was utilised to define LD for input into MAGMA. Genic boundaries were extended to capture regulatory variation, with both conservative and liberal upstream and downstream boundary definition implemented. An extension of 5 kilobases (kb) upstream of the gene, and 1.5 kb downstream was the conservative construct, whilst a larger 35 kb upstream and 10 kb downstream was the liberal construct. Boundaries were longer upstream of the gene in both instances to capture more promoter related variation, as is usual practice^53–55^

Genic *P*-values were transformed to Z-scores with the probit function for input into the gene-set association model. Competitive gene-set association was undertaken by a linear regression model whereby genic Z-scores are the outcome and confounders including gene size and genic minor allele count included as covariates. A one-sided test was performed for the term in the model which specifies whether each gene was within the set of interest (*β*_GS_), such that the null hypothesis is *β*_GS_ = 0 and the alternative *β*_GS_ > 0. When these models are constructed at different *P*_T_, this approach constitutes testing whether the gene-set is more associated than the other genes, for which test-statistics were calculated only including SNPs below the threshold. We defined gene-sets with known drug targets by sourcing hallmark and canonical (BioCarta, KEGG, PID, and Reactome) from the Molecular signatures database (MSigDB)^56^, and retaining those with at least one gene with a high confidence interaction with at least one approved pharmacological agent (T_Clin_ genes), as annotated using the Target Central Resource Database (TCRD v6.1, N_Genes_ = 613)^57^.

### PES candidate gene-set drug repurposing

We tested each candidate PES gene-set for overrepresentation of DrugBank compound targets using WebGestaltR v0.4.2^58^. Compounds were retained for each pathway if they survived FDR correction (*q*<0.05) and were FDA approved (https://www.accessdata.fda.gov/scripts/cder/daf/index.cfm). We then searched the literature for each of these compounds to prioritise them on the basis of side-effects and prior clinical trial evidence. After excluding compounds with only topical formulations available, drugs were reviewed for lung function related adverse events (including all of dyspnea, abnormal breath sounds, decreased respiratory rate, orthopnea, shallow breathing, respiratory distress, respiratory depression or any other related term), important precautions, black-box warnings or any contraindication that might prohibit the drug use in our study population. These data were obtained for each compound using the following databases: drugs.com, Medscape, SIDER v4.1, and the summaries of each product’s characteristics^59^. We also searched for articles that discussed either an improvement or worsening in the lung functions for each compound. The allowed paediatric age and formulation for each compound were also reviewed. The full list of evaluated compounds is detailed in supplementary table 18, with the ranking criteria also detailed in the supplementary methods.

### The PES model for individuals

We defined the model to calculate PES profiles for individuals as follows (equation four). Consider *j* SNPs for *i* individuals, wherein the SNPs are those physically mapped to genes which are members of a candidate PES gene-set (*m*). Let 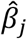, denote the statistical effect size for each variant from the GWAS, multiplied by its dosage *G*_*ij*_. The SNPs included were those below the *P*-value threshold utilised to discover the gene-set.

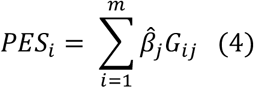

We averaged these scores by the number of SNPs carried by each individual and scaled them using the *scale*() function in R. PES profiles were generated in all instances by first filtering the GWAS summary statistics for common variants (MAF > 0.01) within the genic boundaries of variants which comprise the PES gene-set. The genic boundaries were extended using the liberal or conservative configuration, dependent of which boundary definition was utilised in the gene-set association for that pathway. PRSice v2.2.12 calculated the respective PES, along with genome wide PGS (using the same additive model but genome wide) for FEV_1_ and FVC^60^.

### Lung function PES in the Hunter Community Study cohort

We utilised an independent, genotyped cohort for which spirometry measures were recorded to investigate the phenotype relevance of PES profiles for lung function. Participants were drawn from the Hunter Community Study (HCS), a population-based cohort of individuals aged between 55-85 years, predominantly of European ancestry and residing in Newcastle, New South Wales, Australia. All work was conducted in accordance with ethics committee approvals. Consenting participants completed a series of questionnaires, attended a clinic visit, and provided blood samples. Individuals were recruited by random selection from the New South Wales State electoral roll with detailed recruitment and data collection methods for the HCS described elsewhere^61^. Participants were genotyped using the Affymetrix Axiom Kaiser array and imputed to the Haplotype Reference Consortium (Supplementary Methods)^62^. We retained 2089 unrelated, European ancestry participants and common variants (MAF > 0.01) with high imputation quality (*R*^*2*^ > 0.8). The full description of the imputation and quality control process is provided in in the supplementary methods.

Spirometry data from the HCS was then processed by selecting individuals with non-missing FEV_1_ and FVC. We utilised the maximum FEV_1_ and FVC from four attempts and fitted a linear model which covaried for sex, age, age^2^, height, height^2^, smoking status, self-reported asthma status, and self-reported bronchitis/emphysema status. The phenotype for association testing were residuals from these models transformed via inverse-rank normalisation (Blom transformation) using the RNOmni package. We tested the association between a genome wide PGS for FEV_1_ and FVC (*P*_T_ < 1, 0.5, 0.05, 0.005, 5×10^−5^, 5×10^−8^) with their respective transformed spirometry indices adjusted for the first five SNP derived principal components using PRSice v2.2.12. Similarly, the association between each of the PES profiles with an overrepresentation of FDA-approved drug targets and FEV_1_ and/or FVC were investigated using the same approach. We further adjusted each of these models for genome wide PGS at the same *P*_T_ for which the PES was calculated.

### The relationship between PES and mRNA expression

We obtained RNAseq normalised read counts (PEER normalised RPKM) for 23723 genes which survived QC in the geuvadis dataset (https://www.ebi.ac.uk/arrayexpress/experiments/E-GEUV-1/files/analysis_results/?ref=E-GEUV-1). The geuvadis project performed RNAseq on transformed lymphoblastoid cell lines (LCL) for participants in the 1000 genomes project^21^. We retained 357 European individuals in this dataset for which phase 3 sequencing data was available from the 1000 genomes. The association between normalised mRNA expression for genes part of the candidate gene-set and each PES was tested using a linear model, adjusted for sex, the first three SNP derived principal components, and genome-wide PGS at the same *P*_T_ utilised to calculate the PES. Multiple testing correction was applied for the number of genes in each set via Benjamini-Hochberg method using the *p*.*adjust()* function.

### Transcriptome-wide association studies

A transcriptome-wide association study of each lung function measure was performed using the FUSION software^22^. SNP weights were derived for genes with a significant contribution of *cis* acting SNPs to expression variability (*cis*-*h*^*2*^ *P* < 0.01) using lung and whole blood RNAseq GTEx v7 data (http://gusevlab.org/projects/fusion/). A transcriptome-wide significant gene was defined by accounting for the number of genes with models of genetically regulated expression in lung and whole blood respectively – Lung: *P* < 6.43 × 10^−6^ [α = 0.05/7776], Whole blood: *P* < 8.32 × 10^−6^ [α = 0.05/6007]. We excluded genes within the MHC region due to its LD complexity. Furthermore, we subjected two smoking behaviour phenotypes to TWAS to uncover associations which could be driven by residual effects of smoking. This is inherently conservative as it is possible that genes associated with both lung function and smoking behaviours could exhibit pleiotropic effects, however, as we wish to define drug targets relevant to lung function, the exclusion of these shared genes is warranted. The smoking phenotypes were ‘ever vs never smoked’ and ‘cigarettes smoked per day’ and TWAS was performed using lung and blood for consistency, along with SNP weights from the dorsolateral prefrontal cortex and nucleus accumbens, as these brain regions have been implicated in nicotine addiction. Genes which survived the above were searched using DGidb, with the following criteria utilised to define gene-target pairs, where the drug mode of action matched the sign of the TWAS *Z* value:

i. Tier one – FDA approved compound with at least two lines of evidence for interacting with the target gene,
ii. Tier two – investigational compound (not FDA approved) with at least two lines of evidence for interacting with the target gene.

The TWAS Miami plots were generated using an adapted using an edited version of the TWAS-plotter.V1.0.R script (https://github.com/opain/TWAS-plotter).

### Probabilistic finemapping of druggable TWAS signals

A Bayesian method FOCUS was utilised to finemap TWAS associations which could be therapeutically useful^63^. Given observed TWAS statistics, the marginal posterior inclusion probability (PIP) was calculated and subsequently used to compute a credible set with 90% probability (ρ) of containing the causal gene (c_i_ = 1). As FOCUS allows the null model to be predicted as a possible member of the credible set, we excluded any genes for which that occurred. The credible set (S) was defined by summing normalised PIP such that ρ was exceeded, sorting the genes and then including those genes until at least ρ of the normalized-posterior mass is explained (equation six).

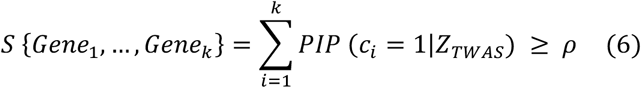

The Bernoulli prior for each causal indicator was set as the default *p* = 1 × 10^−3^, with a default prior variance for effects at causal genes set as 40 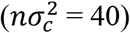. Previous work has demonstrated that FOCUS computed 282s were robust to different specified prior variances^63^, however, we further utilised a more conservative prior of *p* = 1 × 10^−5^ to assess the effect on the *PIP* calculated for candidate druggable genes. In all instances, we utilised a multi-tissue panel obtained from FOCUS GitHub repository which combines GTEx v7 SNP-weights with other FUSION TWAS weights (https://github.com/bogdanlab/focus/wiki,GTEx v7 with METSIM, CMC, YFS, and NTR). The marginal TWAS *Z* to use for finemapping for each locus was selected in the tissue for which the gene was found to be associated via the FUSION TWAS methodology (lung or blood), if available, otherwise by predictive accuracy (cross-validated *R*^2^).

### Host-viral interactome data

We selected three respiratory viruses for which host-viral protein interaction data was previously published: SARS-CoV2, influenza (H1N1), and the human adenovirus (HAdV) family. The host-SARS-CoV2 interactome was defined using affinity-purification mass spectrometry (N_Genes_ = 332, MiST score ≥ 0.7, a SAINTexpress BFDR ≤ 0.05)^25^. We selected 91 proteins which both interact with viral proteins expressed by influenza (mass spectrometry) and siRNA-mediated downregulation reduced viral replication in cultured cells by at least three log_10_ units while retaining >80% cell viability^26^. Finally, the HAdV-host interactome was defined using a protein microarray platform (N_Genes_ = 24), which encompasses 20 viral proteins encoded by five HAdV species^27^. We investigated approved inhibitors or antagonists of these genes using DGidb as described above in the PES candidate gene-set drug repurposing section.

### Overrepresentation of viral prey proteins in glycaemic pathways

The sets of genes which interact with viral proteins for each virus (‘viral prey proteins’), were subjected to overrepresentation analysis using the GENE2FUNC function of FUMA^64^. We selected gene-sets which survived multiple testing correction (*q* < 0.05), which contained at least one of the following key terms related to glycaemic biology: glucose, insulin, diabetes or glucagon. Further, we investigated whether there was a significant overrepresentation of interactions amongst these viral prey proteins overlapping a glycaemic pathway using STRING v11.0^65^. We assembled a list of antidiabetic drug targets by searching compounds annotated with the level two ATC code A10 (Drugs used in diabetes) in DGIdb, retaining drug-gene interactions with two or more lines of evidence (Supplementary Table 59). The interactions between these drug target proteins and the glycaemic SARS-CoV2 prey proteins were investigated once more using STRING, with only interactions scoring > 0.75 considered.

### Code and data availability

All data are publicly available from the references described in the manuscript. Code related to this study can be found at the following link: https://github.com/Williamreay/Lung_function_drug_repurposing_manuscript

## ACKNOWLEDGEMENTS

This study was supported by an NHMRC project grant (1147644). M.J.C. is supported by an NHMRC Senior Research Fellowship (1121474).

Hunter Cohort Study: The research on which this paper is based was conducted as part of the Hunter Community Study, The University of Newcastle. We are grateful to The University of Newcastle for funding and to the men and women of the Hunter region who provided the information recorded.

## AUTHOR CONTRIBUTIONS

W.R.R conceived and designed the study, with input from M.J.C. W.R.R. performed the primary analyses. S.I.E.S. prioritised the drug repurposing candidates. M.P.G assisted with the *pharmagenic enrichment score* analyses. C.R, E.G.H, and R.J.S performed the HCS cohort SNP genotyping, initial data cleaning, and imputation. S.H, M.A.M, R.P, and J.R.A were involved in the collection of the HCS cohort measures. W.R.R. and M.J.C wrote the first draft of the manuscript. All authors contributed to the critical interpretation of the results and preparation of the final manuscript.

## REFERENCES

1. Vasquez, M. M., Zhou, M., Hu, C., Martinez, F. D. & Guerra, S. Low Lung Function in Young Adult Life Is Associated with Early Mortality. Am. J. Respir. Crit. Care Med. 195, 1399–1401 (2017).

2. Young, R. P., Hopkins, R. & Eaton, T. E. Forced expiratory volume in one second: not just a lung function test but a marker of premature death from all causes. Eur. Respir. J. 30, 616–622 (2007).

3. Quaderi, S. A. & Hurst, J. R. The unmet global burden of COPD. Glob Health Epidemiol Genom 3, e4 (2018).

4. Pitocco, D. et al.. The diabetic lung--a new target organ? Rev Diabet Stud 9, 23–35 (2012).

5. Walter, R. E., Beiser, A., Givelber, R. J., O’Connor, G. T. & Gottlieb, D. J. Association between glycemic state and lung function: the Framingham Heart Study. Am. J. Respir. Crit. Care Med. 167, 911–916 (2003).

6. Alonso-Gonzalez, R. et al.. Abnormal lung function in adults with congenital heart disease: prevalence, relation to cardiac anatomy, and association with survival. Circulation 127, 882–890 (2013).

7. Ji, X.-Q., Ji, Y.-B., Wang, S.-X., Zhang, C.-Q. & Lu, D.-G. Alterations of pulmonary function in patients with inflammatory bowel diseases. Ann Thorac Med 11, 249–253 (2016).

8. Yilmaz, A. et al.. Pulmonary involvement in inflammatory bowel disease. World J. Gastroenterol. 16, 4952–4957 (2010).

9. Palmer, L. J. et al.. Familial aggregation and heritability of adult lung function: results from the Busselton Health Study. Eur. Respir. J. 17, 696–702 (2001).

10. Ingebrigtsen, T. S. et al.. Genetic influences on pulmonary function: a large sample twin study. Lung 189, 323–330 (2011).

11. Shrine, N. et al.. New genetic signals for lung function highlight pathways and chronic obstructive pulmonary disease associations across multiple ancestries. Nat. Genet. 51, 481–493 (2019).

12. Reay, W. R., Atkins, J. R., Carr, V. J., Green, M. J. & Cairns, M. J. Pharmacological enrichment of polygenic risk for precision medicine in complex disorders. Sci Rep 10, 879 (2020).

13. Bulik-Sullivan, B. et al. An atlas of genetic correlations across human diseases and traits. Nat. Genet. 47, 1236–1241 (2015).

14. Zheng, J. et al. LD Hub: a centralized database and web interface to perform LD score regression that maximizes the potential of summary level GWAS data for SNP heritability and genetic correlation analysis. Bioinformatics 33, 272–279 (2017).

15. O’Connor, L. J. & Price, A. L. Distinguishing genetic correlation from causation across 52 diseases and complex traits. Nat. Genet. 50, 1728–1734 (2018).

16. Bowden, J. et al. A framework for the investigation of pleiotropy in two-sample summary data Mendelian randomization. Stat Med 36, 1783–1802 (2017).

17. Bowden, J., Davey Smith, G., Haycock, P. C. & Burgess, S. Consistent Estimation in Mendelian Randomization with Some Invalid Instruments Using a Weighted Median Estimator. Genet. Epidemiol. 40, 304–314 (2016).

18. Bowden, J., Davey Smith, G. & Burgess, S. Mendelian randomization with invalid instruments: effect estimation and bias detection through Egger regression. Int J Epidemiol 44, 512–525 (2015).

19. Verbanck, M., Chen, C.-Y., Neale, B. & Do, R. Detection of widespread horizontal pleiotropy in causal relationships inferred from Mendelian randomization between complex traits and diseases. Nat. Genet. 50, 693–698 (2018).

20. Burgess, S., Bowden, J., Fall, T., Ingelsson, E. & Thompson, S. G. Sensitivity Analyses for Robust Causal Inference from Mendelian Randomization Analyses with Multiple Genetic Variants. Epidemiology 28, 30–42 (2017).

21. Lappalainen, T. et al. Transcriptome and genome sequencing uncovers functional variation in humans. Nature 501, 506–511 (2013).

22. Gusev, A. et al. Integrative approaches for large-scale transcriptome-wide association studies. Nat. Genet. 48, 245–252 (2016).

23. Goldstein, R. Z. & Volkow, N. D. Dysfunction of the prefrontal cortex in addiction: neuroimaging findings and clinical implications. Nat. Rev. Neurosci. 12, 652–669 (2011).

24. Scofield, M. D. et al. The Nucleus Accumbens: Mechanisms of Addiction across Drug Classes Reflect the Importance of Glutamate Homeostasis. Pharmacol. Rev. 68, 816–871 (2016).

25. Gordon, D. E. et al. A SARS-CoV-2 protein interaction map reveals targets for drug repurposing. Nature (2020) doi:10.1038/s41586-020-2286-9.

26. Watanabe, T. et al. Influenza virus-host interactome screen as a platform for antiviral drug development. Cell Host Microbe 16, 795–805 (2014).

27. Martinez-Martin, N. et al. The extracellular interactome of the human adenovirus family reveals diverse strategies for immunomodulation. Nat Commun 7, 11473 (2016).

28. Kumar, A. et al. Is diabetes mellitus associated with mortality and severity of COVID-19? A meta-analysis. Diabetes Metab Syndr 14, 535–545 (2020).

29. Davis, W. A. et al. Glycemic exposure is associated with reduced pulmonary function in type 2 diabetes: the Fremantle Diabetes Study. Diabetes Care 27, 752–757 (2004).

30. Gutiérrez-Carrasquilla, L. et al. Effect of Glucose Improvement on Spirometric Maneuvers in Patients With Type 2 Diabetes: The Sweet Breath Study. Diabetes Care 42, 617–624 (2019).

31. van den Borst, B., Gosker, H. R., Zeegers, M. P. & Schols, A. M. W. J. Pulmonary function in diabetes: a metaanalysis. Chest 138, 393–406 (2010).

32. McKeever, T. M., Weston, P. J., Hubbard, R. & Fogarty, A. Lung function and glucose metabolism: an analysis of data from the Third National Health and Nutrition Examination Survey. Am. J. Epidemiol. 161, 546–556 (2005).

33. Barrett-Connor, E. & Frette, C. NIDDM, impaired glucose tolerance, and pulmonary function in older adults. The Rancho Bernardo Study. Diabetes Care 19, 1441–1444 (1996).

34. Talakatta, G. et al. Diabetes induces fibrotic changes in the lung through the activation of TGF-β signaling pathways. Sci Rep 8, 11920 (2018).

35. Mohanty, P. et al. Glucose challenge stimulates reactive oxygen species (ROS) generation by leucocytes. J. Clin. Endocrinol. Metab. 85, 2970–2973 (2000).

36. Sun, Q., Li, J. & Gao, F. New insights into insulin: The anti-inflammatory effect and its clinical relevance. World J Diabetes 5, 89–96 (2014).

37. Lepper, P. M. et al. Serum glucose levels for predicting death in patients admitted to hospital for community acquired pneumonia: prospective cohort study. BMJ 344, e3397 (2012).

38. Jensen, A. V. et al. The impact of blood glucose on community-acquired pneumonia: a retrospective cohort study. ERJ Open Res 3, (2017).

39. Kornum, J. B. et al. Type 2 diabetes and pneumonia outcomes: a population-based cohort study. Diabetes Care 30, 2251–2257 (2007).

40. McAlister, F. A. et al. The relation between hyperglycemia and outcomes in 2,471 patients admitted to the hospital with community-acquired pneumonia. Diabetes Care 28, 810–815 (2005).

41. Wang, Q. et al. O -GlcNAc transferase promotes influenza A virus–induced cytokine storm by targeting interferon regulatory factor–5. Sci. Adv. 6, eaaz7086 (2020).

42. Okayama, H. et al. Bronchodilating effect of intravenous magnesium sulfate in bronchial asthma. JAMA 257, 1076–1078 (1987).

43. Hossein, S. et al. The effect of nebulized magnesium sulfate in the treatment of moderate to severe asthma attacks: a randomized clinical trial. Am J Emerg Med 34, 883–886 (2016).

44. Gu, Z., Eils, R. & Schlesner, M. Complex heatmaps reveal patterns and correlations in multidimensional genomic data. Bioinformatics 32, 2847–2849 (2016).

45. VanderWeele, T. J., Tchetgen Tchetgen, E. J., Cornelis, M. & Kraft, P. Methodological challenges in mendelian randomization. Epidemiology 25, 427–435 (2014).

46. Manning, A. K. et al. A genome-wide approach accounting for body mass index identifies genetic variants influencing fasting glycemic traits and insulin resistance. Nat. Genet. 44, 659–669 (2012).

47. Scott, R. A. et al. Large-scale association analyses identify new loci influencing glycemic traits and provide insight into the underlying biological pathways. Nat. Genet. 44, 991–1005 (2012).

48. Burgess, S., Butterworth, A. & Thompson, S. G. Mendelian randomization analysis with multiple genetic variants using summarized data. Genet. Epidemiol. 37, 658–665 (2013).

49. Slob, E. A. W. & Burgess, S. A comparison of robust Mendelian randomization methods using summary data. Genetic Epidemiology 44, 313–329 (2020).

50. Watanabe, K. et al. A global overview of pleiotropy and genetic architecture in complex traits. Nat. Genet. 51, 1339–1348 (2019).

51. Liu, M. et al. Association studies of up to 1.2 million individuals yield new insights into the genetic etiology of tobacco and alcohol use. Nat. Genet. 51, 237–244 (2019).

52. de Leeuw, C. A., Mooij, J. M., Heskes, T. & Posthuma, D. MAGMA: generalized gene-set analysis of GWAS data. PLoS Comput. Biol. 11, e1004219 (2015).

53. Wray, N. R. et al. Genome-wide association analyses identify 44 risk variants and refine the genetic architecture of major depression. Nat. Genet. 50, 668–681 (2018).

54. Kunkle, B. W. et al. Genetic meta-analysis of diagnosed Alzheimer’s disease identifies new risk loci and implicates Aβ, tau, immunity and lipid processing. Nat. Genet. 51, 414–430 (2019).

55. Reay, W. R. & Cairns, M. J. Pairwise common variant meta-analyses of schizophrenia with other psychiatric disorders reveals shared and distinct gene and geneset associations. Transl Psychiatry 10, 134 (2020).

56. Liberzon, A. et al. The Molecular Signatures Database (MSigDB) hallmark gene set collection. Cell Syst 1, 417–425 (2015).

57. Oprea, T. I. et al. Unexplored therapeutic opportunities in the human genome. Nat Rev Drug Discov 17, 317–332 (2018).

58. Liao, Y., Wang, J., Jaehnig, E. J., Shi, Z. & Zhang, B. WebGestalt 2019: gene set analysis toolkit with revamped UIs and APIs. Nucleic Acids Res. 47, W199–W205 (2019).

59. Kuhn, M., Letunic, I., Jensen, L. J. & Bork, P. The SIDER database of drugs and side effects. Nucleic Acids Res 44, D1075–D1079 (2016).

60. Choi, S. W. & O’Reilly, P. F. PRSice-2: Polygenic Risk Score software for biobanksscale data. GigaScience 8, giz082 (2019).

61. McEvoy, M. et al. Cohort Profile: The Hunter Community Study. International Journal of Epidemiology 39, 1452–1463 (2010).

62. Loh, P.-R. et al. Reference-based phasing using the Haplotype Reference Consortium panel. Nat Genet 48, 1443–1448 (2016).

63. Mancuso, N. et al. Probabilistic fine-mapping of transcriptome-wide association studies. Nat. Genet. 51, 675–682 (2019).

64. Watanabe, K., Taskesen, E., van Bochoven, A. & Posthuma, D. Functional mapping and annotation of genetic associations with FUMA. Nat Commun 8, 1826 (2017).

65. Szklarczyk, D. et al. STRING v11: protein-protein association networks with increased coverage, supporting functional discovery in genome-wide experimental datasets. Nucleic Acids Res. 47, D607–D613 (2019).

